# Hospital-treated infectious diseases, infection burden, and subsequent risk of demyelinating diseases: a prospective cohort study

**DOI:** 10.64898/2026.10.23.26366371

**Authors:** Qingmei Chen, Yueping Shen, Ying Chen, Jianian Hua

**Affiliations:** Department of Neurology, The First Affiliated Hospital of Soochow University, Suzhou, Jiangsu, China; Department of Physical Medicine &Rehabilitation, The First Affiliated Hospital of Soochow University, Suzhou, 215000, Jiangsu Province, China; Department of Epidemiology and Biostatistics, School of Public Health, Medical College of Soochow University, 199 Ren’ai Road, Suzhou, 215123, China

**Keywords:** infection, demyelinating diseases, multiple sclerosis, risk factor

## Abstract

**Background and Objectives:** Epstein–Barr virus infection is an established risk factor for multiple sclerosis (MS). However, associations between a wider range of infections and MS remain unclear, particularly among middle-aged and older adults. Risk factors for other (non-MS) demyelinating diseases are also poorly understood. We examined whether hospital-treated infectious diseases and infection burden were associated with subsequent risk of demyelinating diseases.

**Methods:** This prospective cohort study included 484,292 UK Biobank participants (mean age 56.5 [SD 8.1] years; 54.3% women). Participants with demyelinating diseases at baseline were excluded. Hospital-treated infectious diseases and incident demyelinating diseases were ascertained from linked inpatient records. The primary outcome was any incident demyelinating disease; secondary outcomes were MS and other demyelinating diseases (not mutually exclusive). Infection burden was assessed by episode count, maximum number of concurrent infections, and number of affected anatomical sites. Cox proportional hazards models estimated hazard ratios (HRs) and 95% confidence intervals (CIs), adjusted for sociodemographic, lifestyle, and clinical factors.

**Results:** During a median follow-up of 12.7 years (IQR, 12.0–13.4), 592 participants (0.12%) developed demyelinating diseases, including 436 with MS and 194 with other demyelinating diseases. Hospital-treated infectious diseases were associated with higher risks of all demyelinating diseases (HR 1.80, 95% CI 1.49–2.17), MS (HR 1.88, 95% CI 1.51–2.35), and other demyelinating diseases (HR 1.65, 95% CI 1.18–2.31). For all demyelinating diseases, greater infection burden across all 3 measures was associated with progressively higher risk (all *P* for trend <0.001); HRs were 1.45 (95% CI 1.16–1.81) for 1 infection episode and 2.97 (95% CI 2.22–3.97) for ≥2 episodes versus none.

**Discussion:** Hospital-treated infectious diseases and greater infection burden were associated with higher subsequent risk of demyelinating diseases among middle-aged and older adults. These findings broaden current understanding of infection-related risk and may inform future prevention research.

## Introduction

Demyelinating diseases of the central nervous system (CNS) can cause severe and lasting neurological disability. Multiple sclerosis (MS), the most common type of demyelinating disease, affects more than 150,000 people in the UK, with approximately 7,100 new diagnoses annually.^[1, 2]^ Other demyelinating diseases, such as neuromyelitis optica spectrum disorder (NMOSD), acute transverse myelitis, and clinically isolated syndrome (CIS), can also result in poor functional outcomes.^[3]^ Furthermore, a substantial proportion of individuals presenting with CIS subsequently develop MS.^[4]^ Therefore, understanding risk factors for demyelinating diseases is needed to inform prevention strategies and improve early identification of individuals at increased risk.

Unlike cardiovascular disease, which often be sloganed by reducing cholesterol to prevent, prevention strategies for demyelinating diseases remain less clearly defined.^[5, 6]^ Previous studies have predominantly focused on MS and have reported associations of genetic susceptibility and environmental and lifestyle factors,^[7–10]^ including smoking, body mass index (BMI), educational attainment, and vitamin D levels, with disease risk.^[11–13]^ Epstein–Barr virus (EBV) infection is viewed as a strong risk factor for MS, potentially through its effects on immune pathways.^[14, 15]^ However, the potential contribution of hospital-treated infections, which include a wide range of infections beyond EBV infection and may reflect greater clinical severity than infections managed solely in primary care, remains less studied. Also, immunotherapy for patients with demyelinating disease can increase infection risk, highlighting the importance of carefully assessing the association.^[16–18]^

To our knowledge, only one population-based study reported that hospital-diagnosed infections during adolescence (ages 11–19 years) were associated with a 33% higher risk of MS diagnosis from age 20 years onwards (HR, 1.33).^[19]^ However, these findings were derived from a younger population, and their generalizability to middle-aged and older adults remains unclear. Compared with younger-onset MS, later-onset MS has a different clinical profile, immune function, environmental risk factors, and more rapid disability accumulation,^[20–22]^ which would lead to different results. Moreover, the association between hospital-treated infections and non-MS demyelinating diseases is unknown.

Hence, using data from UK Biobank, a large prospective cohort, we aimed to examine the associations between hospital-treated infectious diseases and the subsequent risk of CNS demyelinating diseases among middle-aged and older adults. We evaluated all demyelinating diseases, MS, and other (non-MS) demyelinating diseases. Then, we examined whether greater infection burden was associated with higher risk. Finally, we explored the potential moderating effects of sociodemographic characteristics, lifestyle factors, and comorbidities on the studied associations.

## Methods

### Study population

This prospective cohort study used data from UK Biobank, which recruited half a million participants aged 37–72 years between 2006 and 2010 across 22 assessment centres in England, Scotland, and Wales. At baseline, participants completed touchscreen questionnaires, participated in interviews and underwent physical and laboratory examinations. A detailed description of the cohort has been published elsewhere.^[23]^ Subsequent health outcomes were ascertained through linkage to electronic health records. UK Biobank received approval from the North West – Haydock Research Ethics Committee (21/NW/0157), and all participants provided written informed consent.

A total of 502,402 participants were initially assessed. Participants who had withdrawn from the study (n = 158), had prevalent demyelinating diseases at baseline (n = 2,113), or had missing infection dates (n = 2) or covariate data (n = 15,837) were excluded, leaving 484,292 participants for analysis (Figure S1).

### Definition of outcomes

The primary outcome was the first incident diagnosis of any CNS demyelinating disease. Secondary outcomes were the first incident diagnoses of MS and other (non-MS) demyelinating diseases.

Participants diagnosed with both MS and other demyelinating diseases could be included in both outcomes. Incident events during follow-up were identified from hospital inpatient records (Data-Fields 41270 and 41271) using the International Classification of Diseases, 10th Revision (ICD-10) and ICD-9 codes. The inpatient records were sourced from Hospital Episode Statistics for England, the Scottish Morbidity Record for Scotland, and the Patient Episode Database for Wales. The earliest diagnosis date was used as the event date. Follow-up time was calculated from baseline recruitment to the date of the event, loss to follow-up, death, or the censoring date (November 2021), whichever came first. Records of demyelinating diseases at baseline were identified from hospital inpatient records and self-reported information (Data-Field 20002). The diagnostic codes are provided in Supplementary Table S1.

### Exposure

The exposure in the main analysis was hospital-treated infectious diseases identified using ICD-10 codes, with the date of the first record of any infection used as the exposure date.^[24]^ Mild upper respiratory tract infections were not included. In analyses by infection type and site, infections were categorised by type (bacterial and viral infections and their subcategories, including bacterial infections with or without sepsis, Gram-positive and Gram-negative bacterial infections, and acute viral infections) and site (including respiratory, urinary, gastrointestinal, and skin infections). Table S2 provides the codes for these infection categories. Participants with a recorded infection diagnosis before baseline were considered exposed from study entry. For participants who were first diagnosed with an infection after baseline, their exposure status changed: they were considered unexposed from study entry until the infection diagnosis and exposed from the time of infection until the end of follow-up.

### Covariates

Covariates were selected based on prior literature and clinical considerations regarding their potential roles in confounding the association between hospital-treated infections and demyelinating diseases,^[5, 13, 25]^ including age, sex, education, Townsend Deprivation Index (TDI), ethnicity, lifestyle factors (smoking status, alcohol consumption, physical activity, and body mass index [BMI]), and comorbidities (hypertension, diabetes, cardiovascular diseases [CVD], and cancer). Comprehensive descriptions of each variable are provided in Tables S3 and S4.

### Statistical analysis

Baseline characteristics of the study population were summarized according to infection and demyelinating disease. Categorical variables are presented as numbers and percentages, and continuous variables as means and standard deviations (SDs). Baseline characteristics were compared using analysis of variance (ANOVA) for continuous variables and chi-square tests for categorical variables. The Kaplan–Meier (KM) method was used to estimate cumulative incidence curves for demyelinating diseases, and log-rank tests were used to compare curves between participants with and without hospital-treated infections. Cox proportional hazards regression models were used to examine the associations of hospital-treated infections with the subsequent risk of demyelinating diseases. Results are reported as hazard ratios (HRs) and 95% confidence intervals (CIs). Two progressively adjusted models were constructed. Model 1 was adjusted for age, sex, educational attainment, and TDI. Model 2 was additionally adjusted for ethnicity, smoking, alcohol consumption, physical activity, BMI, hypertension, diabetes, cancer, and CVD. Analyses were performed separately for demyelinating diseases, MS, and other demyelinating diseases. The proportional hazards assumption was assessed using tests based on Schoenfeld residuals, with no evidence of violation.

We then examined associations according to type and site of infection to assess whether the observed associations were consistent across different categories of hospital-treated infections. Associations for several infection types and sites listed in Table S2 were not estimated due to insufficient numbers of events. Infection burden was assessed using three measures: the number of infection episodes, the maximum number of concurrent infection diagnoses, and the number of affected infection sites.

Infection episodes were defined by the number of recorded infection diagnosis dates, with multiple infection diagnoses recorded on the same date counted as one episode.^[24]^ The maximum number of concurrent infection diagnoses was defined as the largest number of distinct ICD-10 codes recorded on the same date. The number of affected infection sites was defined as the cumulative number of anatomical site categories. Each measure was categorised as 0, 1, or ≥2 and included as a time-varying exposure variable in the Cox analyses, in which the value was updated when an additional infection diagnosis was recorded. Participants with no infection served as the reference group. *P* values for linear trends were assessed by modelling the ordinal categories as continuous variables.

To examine whether the associations observed in the primary analyses differed according to sociodemographic characteristics, educational attainment, lifestyle factors, and comorbidities, subgroup analyses were conducted according to baseline age (<60 or ≥60 years), sex, ethnicity, education, physical activity, smoking, alcohol consumption, BMI, hypertension, diabetes, cancer, or CVD. Potential effect modification was assessed by adding an interaction term of infection × each subgroup variable in the Cox regression models.

Several sensitivity analyses were conducted to assess the robustness of our primary findings. First, analyses were restricted to participants aged ≥50 years at baseline to assess the associations among older adults, using an age cutoff consistent with the definition of late-onset MS.^[22]^ Second, follow-up was censored on December 31, 2019, to minimise the potential influence of the COVID-19 pandemic on both hospital admissions and demyelinating diseases.^[26]^ Third, Fine and Gray models were fitted with death treated as a competing event to account for the possibility that death could preclude the occurrence of demyelinating diseases during follow-up.^[27]^ Fourth, to address missing covariate data, the primary analyses were repeated using multiple imputation. Twenty imputed datasets were generated with 20 iterations, and the estimates from Cox proportional hazards models across the imputed datasets were pooled according to Rubin’s rules.^[28]^ Fifth, given the long prodromal period of MS and the possibility of reverse causation, we repeated the analyses after excluding all participants with hospital-treated infections occurring within 3 or 5 years before baseline.^[29]^ We additionally applied lag periods of 3, 5, and 10 years after infection,^[30]^ during which time exposure was inactive (set to be 0). Sixth, we calculated the E-value, which represents the minimum strength of association that an unmeasured confounder would need to have with both the exposure and outcome, conditional on all measured covariates, to fully explain the observed association.^[31]^

All statistical tests were two-sided, with a *P* value <0.05 considered statistically significant. Statistical analyses were performed using SAS version 9.4 and R version 4.6.1.

## Results

### Cohort description

Among the 484,292 participants, the mean (SD) baseline age was 56.5 (8.1) years; 54.3% were women. During a median follow-up of 12.7 years (IQR, 12.0–13.4), 592 participants (0.12%) developed demyelinating diseases. MS and other demyelinating diseases were identified in 436 and 194 participants, respectively. Among participants who developed demyelinating diseases, the mean (SD) baseline age was 54.4 (8.1) years, and the mean (SD) time from baseline to the first diagnosis was 6.6 (3.4) years. Overall, 125,659 participants (25.9%) had a recorded hospital-treated infection before the end of follow-up. The mean (SD) age at infection was 61.0 (11.1) years.

Younger baseline age, female sex, current smoking, and less physical activity were associated with a higher risk of incident demyelinating diseases (Table 1). Participants who developed demyelinating diseases were also less likely to report alcohol consumption ≥3 times/week and to have hypertension, but more likely to have CVD. Similar patterns were observed for MS, although the differences in hypertension and CVD were not statistically significant. Additionally, the proportion of participants of White ethnicity was 4.7 percentage points higher among those who developed MS than among those who did not. Participants who developed MS were also less likely to hold a college or university degree. Participants who developed other demyelinating diseases were younger at baseline, more often women, and more likely to be current smokers and physically inactive than those who did not. The distribution of BMI categories also differed between the two groups. Participant characteristics stratified by hospital-treated infection status are presented in Table S5. Compared with participants without infections, those with infections were older, more often men, and were slightly more likely to be of non-white ethnicity. They were less likely to hold a college or university degree and had higher TDI. They were also more likely to be current smokers, physically inactive, have a BMI ≥30 kg/m², have hypertension, diabetes, CVD and cancer, and were less likely to report alcohol consumption.

**Table 1.** Participant characteristics stratified by outcome.

| Characteristics | Demyelinating diseases |  |  | Multiple sclerosis |  |  | Other demyelinating diseases |  |  |
| --- | --- | --- | --- | --- | --- | --- | --- | --- | --- |
|  | Without<br>(n=483,700) | With<br>(n=592) | P Value | Without<br>(n=483,856) | With<br>(n=436) | P Value | Without<br>(n=484,098) | With<br>(n=194) | P Value |
| Age, years, mean (SD) | 56.5 (8.1) | 54.4 (8.1) | <0.001 | 56.5 (8.1) | 54.6 (8.2) | <0.001 | 56.5 (8.1) | 54.0 (7.7) | <0.001 |
| Female, n (%) | 262,463 (54.3) | 390 (65.9) | <0.001 | 262,563 (54.3) | 290 (66.5) | <0.001 | 262,730 (54.3) | 123 (63.4) | 0.011 |
| White ethnicity, n (%) | 429,771 (88.9) | 537 (90.7) | 0.151 | 429,900 (88.8) | 408 (93.6) | 0.002 | 430,144 (88.9) | 164 (84.5) | 0.056 |
| Below college or university degree, n (%) | 325,528 (67.3) | 417 (70.4) | 0.104 | 325,632 (67.3) | 313 (71.8) | 0.046 | 325,811 (67.3) | 134 (69.1) | 0.599 |
| Townsend Deprivation Index, mean (SD) | -1.4 (3.1) | -1.2 (3.2) | 0.278 | -1.4 (3.1) | -1.3 (3.2) | 0.925 | -1.4 (3.1) | -0.9 (3.2) | 0.087 |
| Alcohol consumption $\geq 3$ times/week, n (%) | 212,204 (43.9) | 213 (36.0) | <0.001 | 212,263 (43.9) | 154 (35.3) | <0.001 | 212,342 (43.9) | 75 (38.7) | 0.144 |
| Current smokers, n (%) | 50,297 (10.4) | 100 (16.9) | <0.001 | 50,325 (10.4) | 72 (16.5) | <0.001 | 50,358 (10.4) | 39 (20.1) | <0.001 |
| Physical inactivity, n (%) | 85,464 (17.7) | 151 (25.5) | <0.001 | 85,499 (17.7) | 116 (26.6) | <0.001 | 85,568 (17.7) | 47 (24.2) | 0.017 |
| Body mass index, kg/m<sup>2</sup>, n (%) |  |  | 0.974 |  |  | 0.561 |  |  | 0.045 |
| <25 | 160,230 (33.1) | 196 (33.1) |  | 160,277 (33.1) | 149 (34.2) |  | 160,369 (33.1) | 57 (29.4) |  |
| 25–<30 | 207,313 (42.9) | 256 (43.2) |  | 207,393 (42.9) | 176 (40.4) |  | 207,469 (42.9) | 100 (51.5) |  |
| $\geq 30$ | 116,157 (24.0) | 140 (23.6) | | 116,186 (24.0) | 111 (25.5) | | 116,260 (24.0) | 37 (19.1) | |
| Hypertension, n (%) | 260,220 (53.8) | 294 (49.7) | 0.044 | 260,298 (53.8) | 216 (49.5) | 0.075 | 260,418 (53.8) | 96 (49.5) | 0.229 |
| Diabetes, n (%) | 29,447 (6.1) | 44 (7.4) | 0.172 | 29,461 (6.1) | 30 (6.9) | 0.489 | 29,476 (6.1) | 15 (7.7) | 0.339 |
| Cardiovascular diseases, n (%) | 19,332 (4.0) | 35 (5.9) | 0.017 | 19,344 (4.0) | 23 (5.3) | 0.174 | 19,354 (4.0) | 13 (6.7) | 0.055 |
| Cancer, n (%) | 44,010 (9.1) | 51 (8.6) | 0.683 | 44,020 (9.1) | 41 (9.4) | 0.824 | 44,049 (9.1) | 12 (6.2) | 0.158 |
| Any hospital-treated infectious diseases | 125,379 (25.9) | 280 (47.3) | <0.001 | 125,448 (25.9) | 211 (48.4) | <0.001 | 125,570 (25.9) | 89 (45.9) | <0.001 |

### Hospital-treated infectious diseases and demyelinating diseases

The KM-estimated cumulative incidence curves for demyelinating diseases separated during follow-up, with higher cumulative incidence in the infection group than in the group without infection (Figure S2). Similar results were observed for multiple sclerosis and other demyelinating diseases (Figures S3-S4). After adjustment for covariates in Model 1, hospital-treated infections were associated with a higher risk of demyelinating diseases (HR, 1.92; 95% CI, 1.59 to 2.32; *P* < 0.001; Figure 1), with slight attenuation after further adjustment in Model 2 (HR, 1.80; 95% CI, 1.49 to 2.17; *P* < 0.001). Associations were also observed for MS (Model 2: HR, 1.88; 95% CI, 1.51 to 2.35; *P* < 0.001) and other demyelinating diseases (Model 2: HR, 1.65; 95% CI, 1.18 to 2.31; *P* = 0.004). Risk of demyelinating diseases by type and site of infection

**Figure 1.**
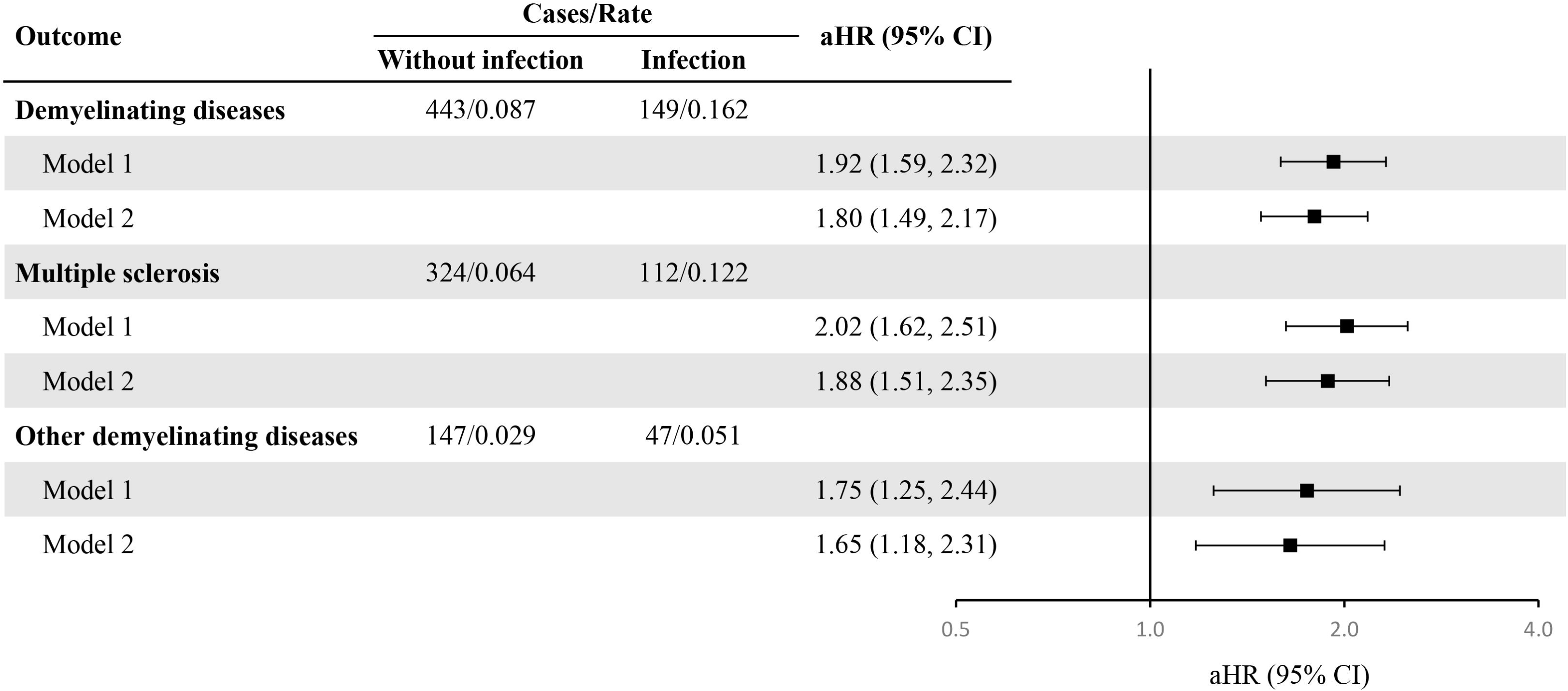
Associations of hospital-treated infectious diseases with the risk of demyelinating diseases. aHRs compare participants with and without hospital-treated infections, with those without infection serving as the reference group. Rates are expressed per 1,000 person-years. Model 1 was adjusted for age, sex, education, and Townsend Deprivation Index. Model 2 was additionally adjusted for ethnicity, smoking, alcohol consumption, physical activity, BMI, hypertension, diabetes, cancer, and cardiovascular disease. Abbreviations: aHR, adjusted hazard ratio; BMI, body mass index.

Positive associations with demyelinating diseases were observed for both bacterial and viral infections and across respiratory, urinary, and gastrointestinal infections, whereas the association with skin infection was not statistically significant in the fully adjusted model (Model 2: HR, 1.47; 95% CI, 0.95–2.28; *P* = 0.086; Figure 2). Similar associations were observed for MS across all infection types and sites, including skin infection (Model 2: HR, 1.77; 95% CI, 1.10–2.86; *P* = 0.019; Figures S5–S6).

**Figure 2.**
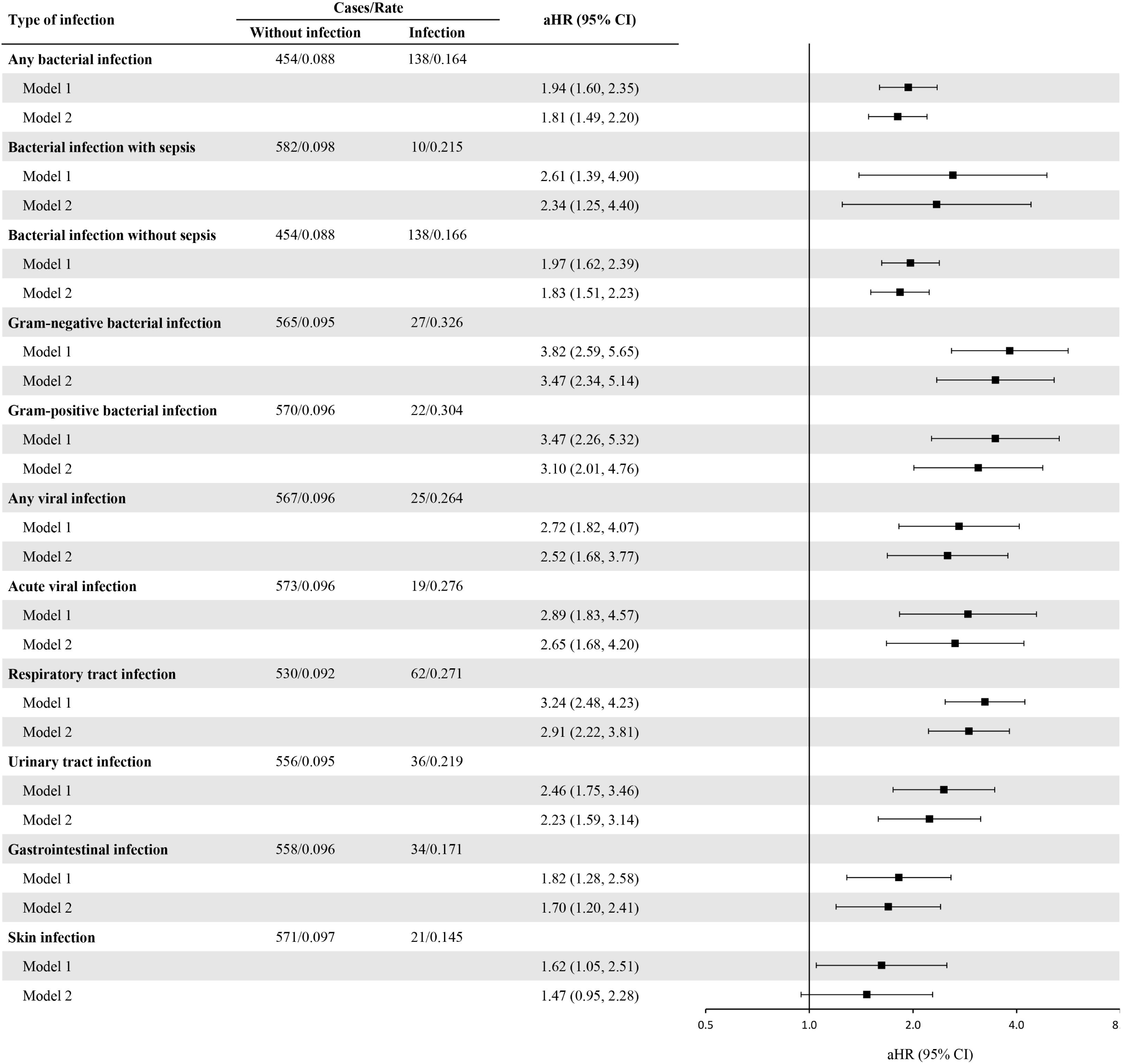
Risk of demyelinating diseases by type and site of infection. For each infection type and site, participants without the corresponding infection served as the reference group. Rates are expressed per 1,000 person-years. Model 1 was adjusted for age, sex, education, and Townsend Deprivation Index. Model 2 was additionally adjusted for ethnicity, smoking, alcohol consumption, physical activity, BMI, hypertension, diabetes, cancer, and cardiovascular disease. Abbreviations: aHR, adjusted hazard ratio; BMI, body mass index.

### Risk of demyelinating diseases by infection burden

Greater infection burden was associated with a progressively higher risk of demyelinating diseases, with consistent patterns observed for the number of infection episodes, maximum number of concurrent infection diagnoses, and number of affected infection sites (all *P* for trend < 0.001; Figure 3). For example, compared with participants without infection, those with a single infection episode had a 45% higher risk of developing demyelinating diseases (Model 2: HR, 1.45; 95% CI, 1.16–1.81), whereas those with at least two infection episodes had a 197% higher risk (Model 2: HR, 2.97; 95% CI, 2.22–3.97). Similar dose-response patterns were also observed for MS and other demyelinating diseases (Figures S7–S8).

**Figure 3.**
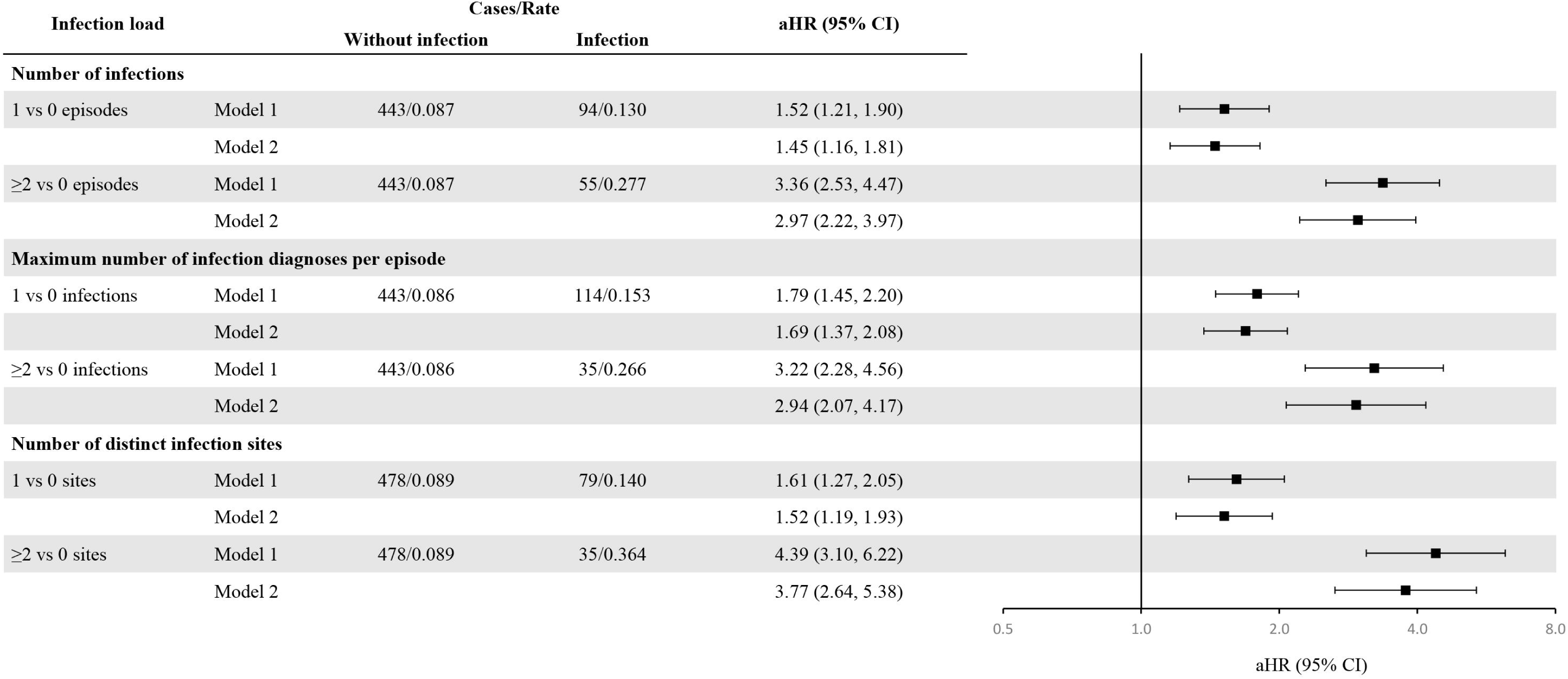
Infection burden and risk of demyelinating diseases. aHRs compare participants in each infection burden category with those without infection. Rates are expressed per 1,000 person-years. Model 1 was adjusted for age, sex, education, and Townsend Deprivation Index. Model 2 was additionally adjusted for ethnicity, smoking, alcohol consumption, physical activity, BMI, hypertension, diabetes, cancer, and cardiovascular disease. Abbreviations: aHR, adjusted hazard ratio; BMI, body mass index.

### Subgroup and sensitivity analyses

The associations between hospital-treated infections and demyelinating diseases were generally consistent across prespecified subgroups (Figure 4). No significant interactions were observed for all demyelinating diseases or MS (all *P* for interaction > 0.05, Figure S9). For other demyelinating diseases, the associations were also generally consistent across subgroups, although heterogeneity was observed across BMI categories (P for interaction = 0.040), with the lowest HR estimate among participants with a BMI <25 kg/m² (Model 2: HR, 0.84; 95% CI, 0.37–1.87; Figure S10).

**Figure 4.**
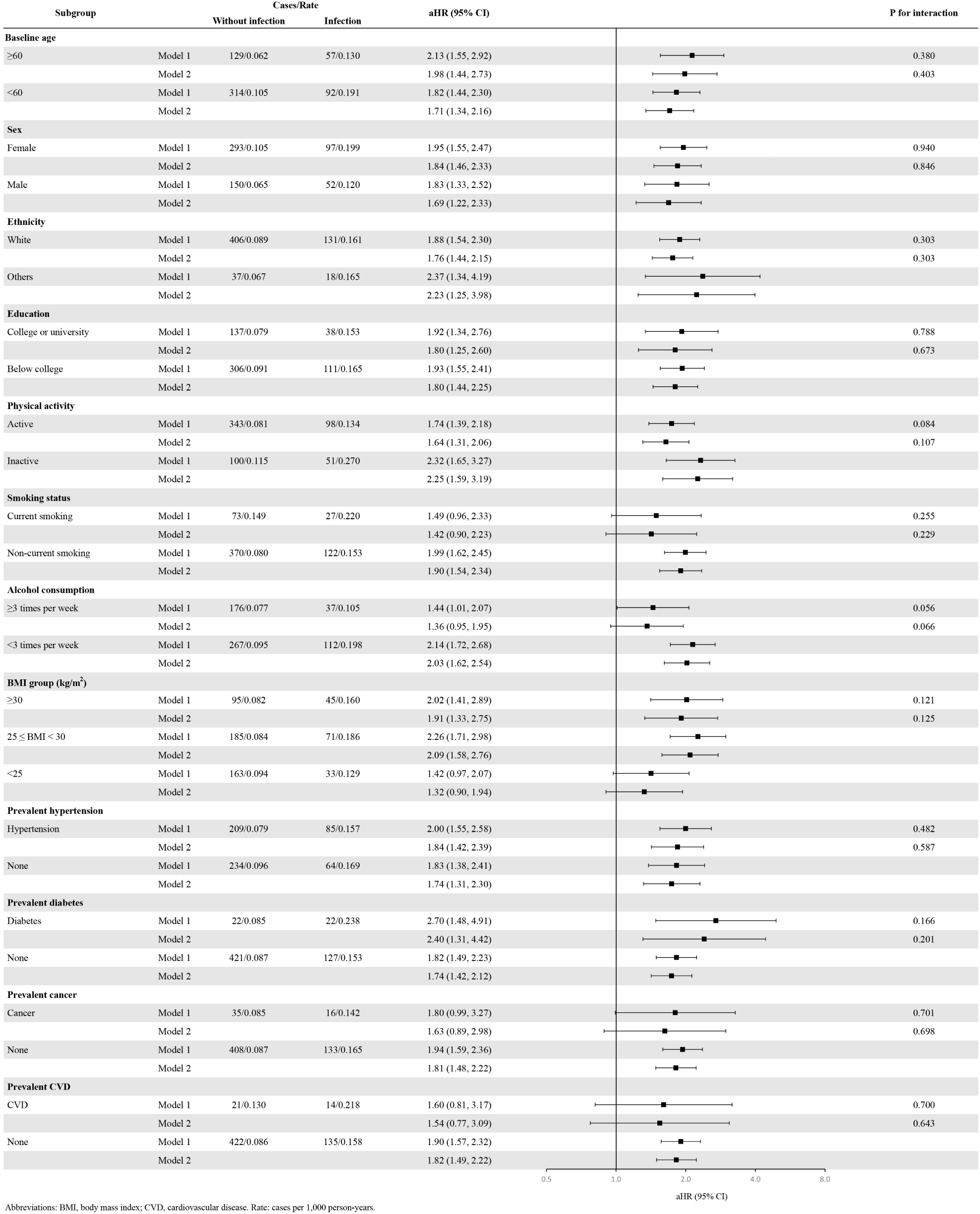
Associations of hospital-treated infections with the risk of demyelinating diseases by baseline characteristics. aHRs compare participants with and without hospital-treated infections, with those without infection serving as the reference group. Rates are expressed per 1,000 person-years. *P* for interaction indicates differences in associations across subgroups. Model 1 was adjusted for age, sex, education, and Townsend Deprivation Index. Model 2 was additionally adjusted for ethnicity, smoking, alcohol consumption, physical activity, BMI, hypertension, diabetes, cancer, and CVD. Abbreviations: aHR, adjusted hazard ratio; BMI, body mass index; CVD, cardiovascular disease.

The primary results were largely unchanged in the following sensitivity analyses: restricting the analysis to participants aged ≥50 years at baseline (Figure S11); censoring follow-up before the COVID-19 pandemic (Figure S12); accounting for the competing risk of death using Fine–Gray models (Figure S13); and using multiple imputation for missing covariates (Figure S14). For other demyelinating diseases, when participants (both cases and controls) with infections occurring within 3 or 5 years before baseline were excluded, the associations became less precise but remained marginally statistically significant (Model 2: HR, 1.75; 95% CI, 1.01–3.04 and HR, 1.87; 95% CI, 1.02–3.43, respectively; Figures S15 and S16). In additional lag analyses, the association with other demyelinating diseases was progressively attenuated as the lag period increased from 3 to 10 years (Model 2: HR, 1.25; 95% CI, 0.84–1.86 for 3 years; HR, 1.19; 95% CI, 0.77–1.83 for 5 years; and HR, 1.09; 95% CI, 0.62–1.93 for 10 years; Figures S17–S19), suggesting that reverse causation might partly contribute to the observed association with other demyelinating diseases. In contrast, the associations for overall demyelinating diseases and MS remained robust across the sensitivity analyses. E-value analyses further supported the robustness of the observed associations to potential unmeasured confounding (Table S6). Notably, the E-values ranged from 5.49 to 7.89 in the group with the highest infection burden for MS, indicating that an unmeasured confounder would need associations of at least 5-fold with both ≥2 infection episodes and MS to fully explain the observed association.

## Discussion

Leveraging a large population-based cohort in the UK, we identified an association between hospital-treated infections and a higher subsequent risk of demyelinating diseases, including MS and other demyelinating diseases. Positive associations were observed for both bacterial and viral infections, for sites of respiratory tract, urinary, and gastrointestinal (except for skin infections), and for various prespecified subgroups. More importantly, greater infection burden was associated with a progressively higher risk of demyelinating diseases. The associations for all demyelinating diseases and MS remained consistent in analyses addressing reverse causation, whereas the association for other demyelinating diseases weakened with increasing lag periods, suggesting that reverse causation may have partly contributed to the observed association.

Prior research has focused mainly on MS, with limited evidence on MS risk factors in middle-aged and older populations (to our knowledge) or on risk factors for non-MS demyelinating diseases.^[26, 32]^ In a Swedish cohort of 2.4 million individuals, hospital-diagnosed infections at ages 11–19 years, but not before age 10 years, were associated with subsequent MS.^[19]^ Another Swedish study reported that pneumonia at ages 11–15 years was associated with approximately twice the odds of subsequent MS.^[29]^ Other studies of MS also highlighted the importance of “exposure timing”. For example, obesity at ages 15–25 years,^[13, 33]^ lower sun exposure before age 15 years,^[11, 34]^ and migration to Denmark before rather than after age 15 years were reported to be associated with an increased risk of MS.^[35]^ Given that the global mean age at MS diagnosis is 32 years and that these studies had relatively limited follow-up (e.g., following participants from age 20 to age 70 would require 50 years of follow-up),^[36]^ their finding may primarily reflect MS risk at younger ages. Our study extends research on risk factors for MS to middle-aged and older populations and also investigates other non-MS demyelinating diseases.

A recent study reported that inpatient-treated infection was associated with disability worsening in patients with MS, compared with outpatient-treated infection.^[17]^ Together with our finding that greater infection burden was associated with an approximately three- to fivefold higher risk of demyelinating diseases, these findings suggest that infection severity and cumulative burden may be relevant to neurological outcomes. Similar observations have been reported in Alzheimer’s disease,^[24]^ in which systemic inflammation was associated with accelerated cognitive decline.^[37]^ Our associations also persisted after adjustment for education, lifestyle factors, and comorbidities and were broadly consistent across subgroups defined by these characteristics, further supporting their robustness. Infections could be a result of demyelinating diseases. High-dose corticosteroids and disease-modifying therapies (DMTs) after the onset of demyelinating diseases increase susceptibility to infections through immunosuppression. In addition, health-related behavioral changes and immune dysregulation during the prodromal phase of MS could potentially lead to infections. To disentangle reverse causation and temporal relation, we conducted sensitivity analyses by imposing a 10-year lag after infection and excluding participants with infections occurring within 5 years before baseline. The results for all demyelinating diseases and MS remained largely unchanged. Although the association with other demyelinating diseases appeared to be partly influenced by reverse causation, this finding should be interpreted cautiously. Acute demyelinating conditions, such as transverse myelitis, MOG antibody-associated disease (MOGAD), and acute disseminated encephalomyelitis (ADEM), often develop 1–4 weeks after an antecedent infection.^[38]^ Such short intervals would be excluded by our sensitivity analyses imposing a lag of at least 3 years. Therefore, the attenuation of the association with other demyelinating diseases in the lag analyses does not necessarily exclude a potential role for infections in triggering non-MS demyelinating diseases.

Using a partly overlapping population, a previous UK Biobank study reported no statistically significant association between hospital-treated infections and MS among participants with diabetes,^[39]^ consistent with our subgroup analysis. The association was statistically significant among participants without diabetes in our study. However, the nonsignificant interaction term provided no evidence that the association differed by diabetes status, and the limited sample size for MS with diabetes might lead to wider confidence intervals that included 1.

For viral infections, EBV remains the leading candidate implicated in MS. Participants with a history of infectious mononucleosis (IM),^[40, 41]^ a common clinical manifestation of EBV infection, had a higher risk of MS (HR = 2.8). Meanwhile, elevated antibody responses to EBV nuclear antigen 1 (EBNA-1) peptides have been observed in patients with MS; increased responses have also been reported in neuromyelitis optica (NMO) and MOGAD, although to a lesser extent than in MS.^[42]^ Other viral infections, including human endogenous retroviruses, measles virus, and rubella virus, have also been investigated in relation to MS.^[43]^ Though our study did not distinguish viral pathogens, the positive association observed in bacterial infections suggests that the association between hospital-treated infections and demyelinating diseases is unlikely to be attributable solely to EBV infection. By using “hospital-treated infectious diseases” as a broader definition of infection, our study identifies a larger population for risk management and provides a clearer and more accessible message for public health promotion.

Several biological mechanisms may underlie the observed association between infections and demyelinating diseases.^[44]^ Infection-induced systemic inflammation may impair blood–brain barrier integrity, activate microglia, and facilitate peripheral immune-cell entry into the central nervous system, potentially promoting neuroinflammation.^[45–47]^ Experimental models of virus-induced demyelination also suggest that infection-related inflammation may contribute to myelin injury through bystander activation, while tissue damage may expose previously sequestered myelin antigens and broaden autoimmune responses through epitope spreading.^[48]^ In MS, immune responses directed against EBNA-1 may trigger CNS autoimmunity via molecular mimicry, supported by evidence that antibodies recognizing EBNA-1 can cross-react with the CNS protein glial cell adhesion molecule.^[15, 47, 49]^ Future experimental studies could clarify whether repeated and concurrent infections contribute to CNS autoimmunity and demyelination.

To our knowledge, this is the first study to report the association between hospital-treated infectious diseases and the subsequent risk of demyelinating diseases. We were also the first to report the association between infection and MS in a middle-aged and older population. Other strengths included the large cohort, long follow-up, adjustment for extensive covariates and addressing reverse causation. Additionally, unlike prior retrospective studies, all participants underwent baseline assessment and were free of demyelinating diseases at baseline, allowing prospective assessment of rare incident outcomes.

However, this study has several limitations. First, UK Biobank’s volunteer cohort (response rate, 5.5%) may not fully represent the entire UK population, although a previous study demonstrated that risk factor associations in the UK Biobank were reproducible in the UK population.^[50]^ Second, although our analyses accounted for multiple confounders and followed participants for a median of 12.7 years, the possibility of unmeasured confounding cannot be ruled out. However, the E-values suggested that an unmeasured confounder would need to be strongly associated with both hospital-treated infections and demyelinating diseases, with associations of approximately threefold for any infection and fivefold for higher infection burden, to fully explain away the observed associations. Few established risk factors for MS have risk estimates of this magnitude. One notable exception is EBV infection: Bjornevik et al. reported a 32-fold increase in MS risk following serum EBV conversion.^[51]^ However, this estimate was specifically derived from participants who were EBV-seronegative at baseline and subsequently seroconverted compared with those who remained seronegative. Only a small proportion of the study population was EBV-seronegative at baseline. On the other hand, the strongest established genetic risk factor for MS, HLA-DRB1*15:01,^[52]^ is associated with an approximately threefold higher risk of MS. Furthermore, we adjusted for a wide range of established environmental and lifestyle risk factors, including BMI and physical activity. Third, reliance on hospital records may have led to underascertainment of undiagnosed or unrecorded cases. Fourth, because of the relatively small number of cases and lack of clinical information, we were unable to further distinguish individual subtypes of other demyelinating diseases. Nevertheless, our results suggested that the effect of infection was similar for MS and other non-MS demyelinating diseases. Fifth, approximately 90% of UK Biobank participants were of White ethnicity. Although positive associations were also observed among participants of other ethnicities in subgroup analyses, the generalizability of our findings to populations with greater ethnic diversity, such as the US population, remains uncertain.

In conclusion, hospital-treated infections were associated with a higher subsequent risk of demyelinating diseases, including MS and other demyelinating diseases, among middle-aged and older adults. Greater infection burden was associated with a progressively higher risk. Our data also reinforce the need for further research into the relationship between infections and other non-MS demyelinating diseases.

## Data Availability

Data are from UK Biobank and are not allowed to be shared.

## Acknowledgement

We thank all participants and staff from the UK Biobank study.

## Glossary

ADEM: acute disseminated encephalomyelitis
BMI: body mass index
CIS: clinically isolated syndrome
CNS: central nervous system
COVID-19: coronavirus disease 2019
CVD: cardiovascular disease
DMTs: disease-modifying therapies
EBNA-1: Epstein–Barr virus nuclear antigen 1
EBV: Epstein–Barr virus
HR: hazard ratio
ICD-10: International Classification of Diseases, Tenth Revision
IM: infectious mononucleosis
MOG: myelin oligodendrocyte glycoprotein
MOGAD: myelin oligodendrocyte glycoprotein antibody-associated disease
MS: multiple sclerosis
NMO: neuromyelitis optica
NMOSD: neuromyelitis optica spectrum disorder
TDI: Townsend Deprivation Index.

## Funding

This work was supported by the following: Boxi Natural Science Foundation (BXQN2024022, Jianian Hua), National Natural Science Foundation of China (82673372, Qingmei Chen) and the Suzhou Basic Research Special Project (SSD2024074, Qingmei Chen).

## Contribution Statement

JH and QMC conceived the study and designed the statistical analyses. YS and JH organized the database. JH and YC did the statistical analyses and prepared the draft of the manuscript. JH and QX substantively revised the manuscript. All authors contributed to the interpretation of data. The corresponding authors attest that all listed authors meet authorship criteria and that no others meeting the criteria have been omitted. All authors read and approved the final manuscript.

## Notes

### Competing Interest Statement

The authors have declared no competing interest.

### Author Declarations

We used data from the UK Biobank, a pulic database but not allowed to share the data.

