## Supplementary Material for "Hospital-treated infectious diseases, infection burden, and subsequent risk of demyelinating diseases: a prospective cohort study"

Supplementary Materials

### Table S1. Codes for demyelinating diseases

| Diseases | ICD-10 | ICD-9 | Data-field 20002 |
| --- | --- | --- | --- |
| Any demyelinating disease | Both codes below | Both codes below | Both codes below |
| Multiple sclerosis (MS) | G35 | 340, 3409 | 1261 |
| Other demyelinating diseases | G36, G360, G361, G368, G369, G37* | 341, 3410, 3411, 3418, 3419 | 1397 |

### Table S2. Codes for hospital-treated infectious diseases

|  | **ICD-10** |
| --- | --- |
| **Any infectious diseases** | ICD-10 BLOCK: A00-A09, A15-A19, A20-A28, A30-A49, A50-A64, A65-A69, A70-A74, A75-A79, A80-A89, A92-A99, B00-B09, B15-B19, B20-B24, B25-B34, B35-B49, B50-B64, B65-B83, B85-B89, B90-B94, B95-B98, B99-B99 Exact codes: C46, D733, E321, G00, G000, G001, G002, G003, G008, G009, G020, G021, G028, G041, G042, G050, G051, G052, H010, H050, H061, H100, H105, H130, H190, H191, H192, H220, H320, H440, H600, H601, H602, H603, H620, H621, H622, H623, H660, H670, H671, H700, H750, I301, I320, I321, I330, I400, I410, I411, I412, I430, I520, I521, I681, J010, J02, J020, J028, J029, J03, J030, J038, J039, J051, J15, J150, J151, J152, J153, J154, J155, J156, J157, J158, J159, J16, J160, J168, J170, J171, J172, J173, J178, J20, J200, J201, J202, J203, J204, J205, J206, J207, J208, J209, J210, J211, J218, J219, J390, J391, J851, J852, J853, K044, K045, K046, K047, K050, K052, K053, K054, K113, K122, K230, K231, K570, K572, K574, K578, K630, K650, K670, K671, K672, K673, K678, K750, K770, L701, M00, M000, M001, M002, M008, M009, M010, M011, M012, M013, M014, M015, M016, M018, M462, M463, M465, M490, M491, M492, M493, M600, M630, M631, M632, M650, M651, M710, M711, M726, M730, M731, M860, M861, M862, M863, M864, M865, M866, M868, M869, N080, N136, N151, N160, N290, N291, N300, N340, N390, N410, N412, N700, N710, N730, N733, N74, N740, N741, N742, N743, N744, N748, N751, N760, N764, N770, N771, O030, O035, O040, O045, O050, O055, O060, O065, O070, O075, O080, O230, O231, O232, O233, O234, O235, O239, O753, O910, O911, O98, O980, O981, O982, O983, O984, O985, O986, O987, O988, O989, P23, P230, P231, P232, P233, P234, P235, P236, P238, P239, P350, P351, P352, P353, P36, P360, P361, P362, P363, P364, P365, P368, P369, P37, P370, P371, P372, P373, P374, P375, P378, P379, P39, P390, P391, P392, P393, P394, P398, P399, R572, R650, R651, Z21 First three characters: G01*, G03*, G06*, G07*, H00*, J04*, J09*, J10*, J11*, J12*, J13*, J14*, J18*, J21*, J22*, J36*, J86*, K02*, K35*, K61*, L00*, L01*, L02*, L03*, L04*, L05*, L08*, M86*, N10*, N45*, N61*, O85*, O86*, P38* |
|  | **Infection type** |
| **Any bacterial infection** | Exact codes: A02, A020, A021, A022, A028, A029, A03, A030, A031, A032, A033, A038, A039, A04, A040, A041, A042, A043, A044, A045, A046, A047, A048, A049, A05, A050, A051, A052, A053, A054, A058, A059, A17, A170, A171, A178, A179, A20, A200, A201, A202, A203, A207, A208, A209, A21, A210, A211, A212, A213, A217, A218, A219, A22, A220, A221, A222, A227, A228, A229, A23, A230, A231, A232, A233, A238, A239, A24, A240, A241, A242, A243, A25, A250, A251, A259, A26, A260, A267, A268, A269, A28, A280, A281, A282, A288, A289, A32, A320, A321, A327, A328, A329, A39, A390, A391, A392, A393, A394, A395, A398, A399, A40, A400, A401, A402, A403, A408, A409, A41, A410, A411, A412, A413, A414, A415, A418, A419, A42, A420, A421, A422, A427, A428, A429, A43, A430, A431, A438, A44, A440, A441, A448, A449, A480, A481, A482, A483, A484, A488, A49, A490, A491, A492, A493, A498, A499, A50, A500, A501, A502, A503, A504, A505, A506, A507, A509, A51, A510, A511, A512, A513, A514, A515, A519, A52, A520, A521, A522, A523, A527, A528, A529, A54, A540, A541, A542, A543, A544, A545, A546, A548, A549, A560, A561, A562, A563, A564, A568, A69, A690, A691, A692, A698, A699, A740, A748, A749, A75, A750, A751, A752, A753, A759, A77, A770, A771, A772, A773, A778, A779, A79, A790, A791, A798, A799, B471, B960, B961, B962, B963, B964, B965, B966, B967, B968, B980, B981, D733, E321, G00, G000, G001, G002, G003, G008, G009, G042, G050, H010, H050, H100, H105, H440, H600, H601, H602, H603, H620, H660, H670, H700, I320, I330, I410, I520, J010, J02, J020, J028, J029, J03, J030, J038, J039, J051, J15, J150, J151, J152, J153, J154, J155, J156, J157, J158, J159, J16, J160, J168, J170, J178, J200, J201, J202, J390, J391, J851, J852, J853, K044, K045, K046, K047, K050, K052, K053, K054, K113, K122, K230, K570, K572, K574, K578, K630, K650, K670, K671, K672, K673, K750, L701, M00, M000, M001, M002, M008, M009, M010, M011, M012, M013, M462, M463, M465, M490, M491, M492, M630, M650, M651, M710, M711, M726, M730, M731, M860, M861, M862, M863, M864, M865, M866, M868, M869, N136, N151, N290, N300, N340, N390, N410, N412, N700, N710, N730, N733, N740, N741, N742, N743, N744, N751, N760, N764, O030, O035, O040, O045, O050, O055, O060, O065, O070, O075, O080, O230, O231, O232, O233, O234, O235, O239, O753, O910, O911, O980, O981, O982, O983, O988, O989, P23, P231, P232, P233, P234, P235, P236, P239, P36, P360, P361, P362, P363, P364, P365, P368, P369, P370, P372, P390, P391, P392, P393, P394 First three characters: A00*, A01*, A09*, A15*, A16*, A18*, A19*, A27*, A30*, A31*, A33*, A34*, A35*, A36*, A37*, A38*, A46*, A53*, A55*, A57*, A58*, A64*, A65*, A66*, A67*, A68*, A70*, A71*, A78*, B95*, G01*, G06*, H00*, J04*, J13*, J14*, J18*, J22*, J36*, J86*, K02*, K35*, K61*, L00*, L01*, L02*, L03*, L04*, L05*, L08*, M86*, N10*, N45*, N61*, O85*, O86*, P38* |
| **Bacterial infection with sepsis** | Exact codes: A021, A207, A217, A227, A327, A391, A392, A393, A394, A40, A400, A401, A402, A403, A408, A409, A41, A410, A411, A412, A413, A414, A415, A418, A419, A427, A440, A483, A484, P36, P360, P361, P362, P363, P364, P365, P368, P369, P372 |
| **Bacterial infection without sepsis** | Exact codes: A02, A020, A022, A028, A029, A03, A030, A031, A032, A033, A038, A039, A04, A040, A041, A042, A043, A044, A045, A046, A047, A048, A049, A05, A050, A051, A052, A053, A054, A058, A059, A17, A170, A171, A178, A179, A20, A200, A201, A202, A203, A208, A209, A21, A210, A211, A212, A213, A218, A219, A22, A220, A221, A222, A228, A229, A23, A230, A231, A232, A233, A238, A239, A24, A240, A241, A243, A25, A250, A251, A259, A26, A260, A267, A268, A269, A28, A280, A281, A282, A288, A289, A32, A320, A321, A328, A329, A39, A390, A395, A398, A399, A42, A420, A421, A422, A428, A429, A43, A430, A431, A438, A44, A441, A448, A449, A480, A481, A482, A488, A49, A490, A491, A492, A493, A498, A499, A50, A500, A501, A502, A503, A504, A505, A506, A507, A509, A51, A510, A511, A512, A513, A514, A515, A519, A52, A520, A521, A522, A523, A527, A528, A529, A54, A540, A541, A542, A543, A544, A545, A546, A548, A549, A560, A561, A562, A563, A564, A568, A69, A690, A691, A692, A698, A699, A740, A748, A749, A75, A750, A751, A752, A753, A759, A77, A770, A771, A772, A773, A778, A779, A79, A790, A791, A798, A799, B471, B960, B961, B962, B963, B964, B965, B966, B967, B968, B980, B981, D733, E321, G00, G000, G001, G002, G003, G008, G009, G042, G050, H010, H050, H100, H105, H440, H600, H601, H602, H603, H620, H660, H670, H700, I320, I330, I410, I520, J010, J02, J020, J028, J029, J03, J030, J038, J039, J051, J15, J150, J151, J152, J153, J154, J155, J156, J157, J158, J159, J16, J160, J168, J170, J178, J200, J201, J202, J390, J391, J851, J852, J853, K044, K045, K046, K047, K050, K052, K053, K054, K113, K122, K230, K570, K572, K574, K578, K630, K650, K670, K671, K672, K673, K750, L701, M00, M000, M001, M002, M008, M009, M010, M011, M012, M013, M462, M463, M465, M490, M491, M492, M630, M650, M651, M710, M711, M726, M730, M731, M860, M861, M862, M863, M864, M865, M866, M868, M869, N136, N151, N290, N300, N340, N390, N410, N412, N700, N710, N730, N733, N740, N741, N742, N743, N744, N751, N760, N764, O030, O035, O040, O045, O050, O055, O060, O065, O070, O075, O080, O230, O231, O232, O233, O234, O235, O239, O753, O910, O911, O980, O981, O982, O983, O988, O989, P23, P231, P232, P233, P234, P235, P236, P239, P370, P390, P391, P392, P393, P394 First three characters: A00*, A01*, A09*, A15*, A16*, A18*, A19*, A27*, A30*, A31*, A33*, A34*, A35*, A36*, A37*, A38*, A46*, A53*, A55*, A57*, A58*, A64*, A65*, A66*, A67*, A68*, A70*, A71*, A78*, B95*, G01*, G06*, H00*, J04*, J13*, J14*, J18*, J22*, J36*, J86*, K02*, K35*, K61*, L00*, L01*, L02*, L03*, L04*, L05*, L08*, M86*, N10*, N45*, N61*, O85*, O86*, P38* |
| **Gram-positive bacterial infection** | Exact codes: A047, A050, A051, A052, A054, A22, A220, A221, A222, A227, A228, A229, A26, A260, A267, A268, A269, A32, A320, A321, A327, A328, A329, A40, A400, A401, A402, A403, A408, A409, A410, A411, A412, A42, A420, A421, A422, A427, A428, A429, A43, A430, A431, A438, A480, A483, A490, A491, B471, B967, G001, G002, G003, J020, J030, J152, J153, J154, J202, M000, M001, M002, P232, P233, P360, P361, P362, P363, P372 First three characters: A33*, A34*, A35*, A36*, A38*, B95*, J13*, L00*, L01* |
| **Gram-negative bacterial infection** | Exact codes: A02, A020, A021, A022, A028, A029, A03, A030, A031, A032, A033, A038, A039, A040, A041, A042, A043, A044, A045, A046, A053, A20, A200, A201, A202, A203, A207, A208, A209, A21, A210, A211, A212, A213, A217, A218, A219, A23, A230, A231, A232, A233, A238, A239, A24, A240, A241, A242, A243, A25, A250, A251, A259, A280, A281, A282, A39, A390, A391, A392, A393, A394, A395, A398, A399, A413, A415, A44, A440, A441, A448, A449, A481, A482, A484, A492, A50, A500, A501, A502, A503, A504, A505, A506, A507, A509, A51, A510, A511, A512, A513, A514, A515, A519, A52, A520, A521, A522, A523, A527, A528, A529, A54, A540, A541, A542, A543, A544, A545, A546, A548, A549, A560, A561, A562, A563, A564, A568, A692, A698, A699, A740, A748, A749, A75, A750, A751, A752, A753, A759, A77, A770, A771, A772, A773, A778, A779, A79, A790, A791, A798, A799, B961, B962, B963, B964, B965, B966, B980, B981, G000, J150, J151, J155, J156, J160, J201, K670, K671, K672, M010, M012, M491, M492, M730, M731, N290, N742, N743, N744, O981, O982, P231, P234, P235, P364 First three characters: A00*, A01*, A27*, A37*, A53*, A55*, A57*, A58*, A65*, A66*, A67*, A68*, A70*, A71*, A78*, J14* |
| **Any viral infection** | Exact codes: A080, A081, A082, A083, A084, A630, A811, A812, A92, A920, A921, A922, A923, A924, A928, A929, A93, A930, A931, A932, A938, A98, A980, A981, A982, A983, A984, A985, A988, B00, B000, B001, B002, B003, B004, B005, B007, B008, B009, B01, B010, B011, B012, B018, B019, B02, B020, B021, B022, B023, B027, B028, B029, B05, B050, B051, B052, B053, B054, B058, B059, B06, B060, B068, B069, B080, B081, B082, B083, B084, B085, B088, B15, B150, B159, B16, B160, B161, B162, B169, B19, B190, B199, B210, B22, B220, B221, B222, B227, B26, B260, B261, B262, B263, B268, B269, B27, B270, B271, B279, B33, B330, B331, B332, B333, B334, B338, B34, B340, B341, B342, B343, B344, B348, B349, B97, B970, B971, B972, B973, B974, B975, B976, B977, B978, C46, G020, G041, G051, H191, H621, H671, I411, J171, J203, J204, J205, J206, J207, J210, J211, J218, J219, M014, M015, O984, O985, O987, P230, P350, P351, P352, P353, Z21 First three characters: A60*, A80*, A83*, A84*, A85*, A86*, A87*, A88*, A89*, A90*, A91*, A94*, A95*, A96*, A97*, A99*, B04*, B07*, B09*, B17*, B18*, B20*, B21*, B23*, B24*, B25*, B30*, J09*, J10*, J11*, J12*, J21* |
| **Herpes virus (persistent) infection** | Exact codes: H191 First three characters: A60*, B00*, B01*, B02* |
| **Acute viral infection** | Exact codes: A080, A081, A082, A083, A084, A92, A920, A921, A922, A923, A924, A928, A929, A93, A930, A931, A932, A938, A98, A980, A981, A982, A983, A984, A985, A988, B05, B050, B051, B052, B053, B054, B058, B059, B06, B060, B068, B069, B080, B081, B083, B084, B085, B088, B15, B150, B159, B16, B160, B161, B162, B169, B19, B190, B199, B26, B260, B261, B262, B263, B268, B269, B33, B330, B331, B332, B334, B338, B34, B340, B341, B342, B343, B348, B349, B97, B970, B971, B972, B974, B975, B976, B978, H671, I411, J171, J203, J204, J205, J206, J207, J210, J211, J218, J219, M014, M015, O985, P230, P350 First three characters: A80*, A83*, A84*, A85*, A86*, A87*, A88*, A89*, A90*, A91*, A94*, A95*, A96*, A97*, A99*, B04*, B07*, B09*, B17*, B30*, J09*, J10*, J11*, J12*, J21* |
| **Other potentially persistent viral infection** | Exact codes: A630, A811, A812, B22, B220, B221, B222, B227, B333, B344, B973, B977, G041, O984, O987, P353, Z21 First three characters: B18*, B20*, B21*, B23*, B24* |
|  | **Infection site** |
| **Respiratory tract infection** | Exact codes: J010, J02, J020, J028, J029, J03, J030, J038, J039, J051, J15, J150, J151, J152, J153, J154, J155, J156, J157, J158, J159, J16, J160, J168, J170, J171, J172, J173, J178, J20, J200, J201, J202, J204, J205, J206, J208, J209, J210, J211, J218, J219, J390, J391, J851, J852, J853, A150, A159, A169, A310, A319, A481, B440, B441 First three characters: J04*, J09*, J10*, J11*, J12*, J13*, J14*, J18*, J21*, J22*, J36*, J86* |
| **Gastrointestinal infection** | Exact codes: K044, K045, K047, K050, K052, K053, K113, K122, K230, K570, K572, K574, K578, K61, K630, K650, K750, K770, A020, A045, A047, A049, A081, A084, A090, A099, B159, B169, B178, B181, B182, B199, B370 First three characters: K35* |
| **Cardiovascular infection** | Exact codes: I301, I320, I321, I330, I400, I410, I411, I412, I430 |
| **Skin infection** | Exact codes: L050, B351, B353, B359 First three characters: L00*, L01*, L02*, L03*, L04*, L08* |
| **Urinary tract infection** | Exact codes: N080, N136, N151, N160, N291, N300, N340, N390 |
| **Genital infection** | Exact codes: N410, N412, N700, N710, N730, N733, N741, N748, N760, N764, N770, N771, A630, B373 First three characters: N45*, N61*, N74* |

*Inclusion of all ICD-10 codes sharing the same first three characters.

### Table S3. Definitions and codes of covariates

| Covariates^*^ | Code | Categories for the current study |
| --- | --- | --- |
| Age | Data-Field 21022 | Continuous |
| Sex | Data-Field 31 | 0 = Female; 1 = Male |
| Ethnicity | Data-Field 21000 | 0 = Other ethnicity; 1 = White ethnicity |
| Education | Data-Field 6138 | 0 = Below college or university degree; 1 = College or university degree |
| Townsend Deprivation Index | Data-Field 189 | Continuous |
| Alcohol consumption ≥3 times/week | Data-Fields 20117 and 1558 | 0 = No; 1 = Yes |
| Current smokers | Data-Field 20116 | 0 = No; 1 = Yes |
| Physical activity | Data-Fields 884 and 904 | 0 = Less than 2 days of moderate or vigorous physical activity per week; 1 = At least 2 days of moderate physical activity per week OR at least 2 days of vigorous physical activity per week |
| BMI | Data-Field 21001 |  |
| BMI < 25 kg/m² |  | 0 = No; 1 = Yes |
| 25 ≤ BMI < 30 kg/m² |  | 0 = No; 1 = Yes |
| BMI ≥ 30 kg/m² |  | 0 = No; 1 = Yes |
| Hypertension | Data-Field 20002, 6150, 6177, 6153, 4080, 4079, 131286 | 0 = No; 1 = Yes |
| Diabetes | Data-Field 20002, 2443, 6177, 6153, 130706, 130708, 130710, 130712, 130714, 132202, 30750; ICD-10 E10–E14 or O24 in Data-Fields 41270/41280 | 0 = No; 1 = Yes |
| CVD | Data-Field 20002, 42000; ICD-10 I21–I23 or I25.2 in Data-Fields 41270/41280  Data-Field 20002, 42006; ICD-10 I60–I64 in Data-Fields 41270/41280 | 0 = No; 1 = Yes |
| Cancer | Data-Field 40005 | 0 = No; 1 = Yes |

Abbreviations: BMI, body mass index; CVD, cardiovascular disease.

^*^ Hypertension was defined by a history of hypertension before baseline, blood pressure ≥140/90 mmHg, or use of antihypertensive medication. Diabetes was defined by a history of diabetes before baseline, HbA1c ≥6.5%, or use of glucose-lowering medication or insulin. CVD was defined by myocardial infarction or stroke.

### Table S4. Missing data for covariates

| Covariates | Number of missing | Percentage (%)* |
| --- | --- | --- |
| Age | 3 | 0.001 |
| Sex | 0 | 0 |
| Ethnicity | 2,751 | 0.55 |
| Education | 0 | 0 |
| Townsend Deprivation Index | 0 | 0 |
| Alcohol consumption ≥3 times/week | 1,635 | 0.33 |
| Current smokers | 2,915 | 0.58 |
| Physical activity | 12,100 | 2.42 |
| BMI | 0 | 0 |
| Hypertension | 0 | 0 |
| Diabetes | 0 | 0 |
| CVD | 0 | 0 |
| Cancer | 0 | 0 |

Abbreviations: BMI, body mass index; CVD, cardiovascular disease.

*Percentage of 500,129 participants before exclusion of missing covariate data.

### Table S5. Participant characteristics stratified by the presence of hospital-treated infectious diseases

|  | **Without infection** | **Infection** | **Total** |  |
| --- | --- | --- | --- | --- |
| **Characteristics** | **(n=358,633)** | **(n=125,659)** | **(n=484,292)** | ***p* value** |
| Age, years, mean (SD) | 55.9 (8.1) | 58.2 (8.0) | 56.5 (8.1) | <0.001 |
| Female, n (%) | 198,580 (55.4) | 64,273 (51.1) | 262,853 (54.3) | <0.001 |
| White ethnicity, n (%) | 318,993 (88.9) | 111,315 (88.6) | 430,308 (88.9) | 0.002 |
| Below college or university degree, n (%) | 233,772 (65.2) | 92,173 (73.4) | 325,945 (67.3) | <0.001 |
| Townsend Deprivation Index, mean (SD) | -1.5 (3.0) | -0.9 (3.3) | -1.4 (3.1) | <0.001 |
| Alcohol consumption ≥3 times/week, n (%) | 162,436 (45.3) | 49,981 (39.8) | 212,417 (43.9) | <0.001 |
| Current smokers, n (%) | 33,107 (9.2) | 17,290 (13.8) | 50,397 (10.4) | <0.001 |
| Physical inactivity, n (%) | 59,983 (16.7) | 25,632 (20.4) | 85,615 (17.7) | <0.001 |
| Body mass index, kg/m², n (%) |  |  |  | <0.001 |
| < 25 | 125,508 (35.0) | 34,918 (27.8) | 160,426 (33.1) |  |
| 25 ≤ BMI < 30 | 155,054 (43.2) | 52,515 (41.8) | 207,569 (42.9) |  |
| ≥ 30 | 78,071 (21.8) | 38,226 (30.4) | 116,297 (24.0) |  |
| Hypertension, n (%) | 183,957 (51.3) | 76,557 (60.9) | 260,514 (53.8) | <0.001 |
| Diabetes, n (%) | 16,474 (4.6) | 13,017 (10.4) | 29,491 (6.1) | <0.001 |
| CVD, n (%) | 10,238 (2.9) | 9,129 (7.3) | 19,367 (4.0) | <0.001 |
| Cancer, n (%) | 28,282 (7.9) | 15,779 (12.6) | 44,061 (9.1) | <0.001 |

Abbreviations: CVD, cardiovascular disease.

### Table S6. E-values of associations between hospital-treated infectious diseases and demyelinating diseases

| Exposure | Outcome | E-value* |
| --- | --- | --- |
| Primary analysis | Demyelinating diseases | 3.00 |
|  | Multiple sclerosis | 3.17 |
|  | Other demyelinating diseases | 2.69 |
| By infection burden |  |  |
| 1 episode | Demyelinating diseases | 2.26 |
|  | Multiple sclerosis | 2.15 |
|  | Other demyelinating diseases | 2.37 |
| ≥2 episodes | Demyelinating diseases | 5.39 |
|  | Multiple sclerosis | 5.79 |
|  | Other demyelinating diseases | 4.98 |
| 1 infection | Demyelinating diseases | 2.77 |
|  | Multiple sclerosis | 2.75 |
|  | Other demyelinating diseases | 2.85 |
| ≥2 infections | Demyelinating diseases | 5.33 |
|  | Multiple sclerosis | 5.49 |
|  | Other demyelinating diseases | 4.86 |
| 1 site | Demyelinating diseases | 2.41 |
|  | Multiple sclerosis | 2.54 |
|  | Other demyelinating diseases | 2.19 |
| ≥2 sites | Demyelinating diseases | 7.00 |
|  | Multiple sclerosis | 7.89 |
|  | Other demyelinating diseases | 4.86 |

*E-values were calculated from results in Model 2.

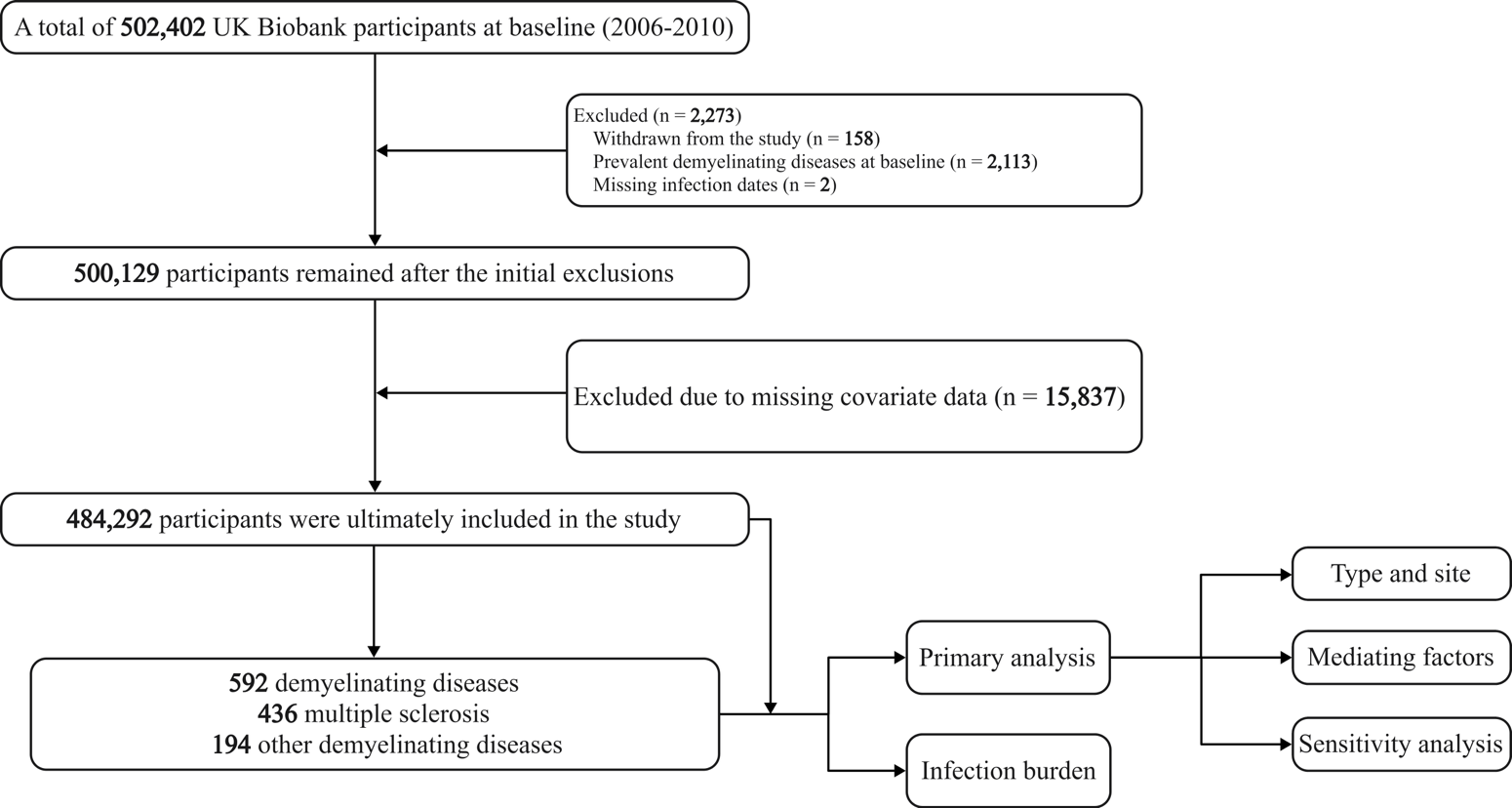

### Figure S1. Flowchart of the study design.

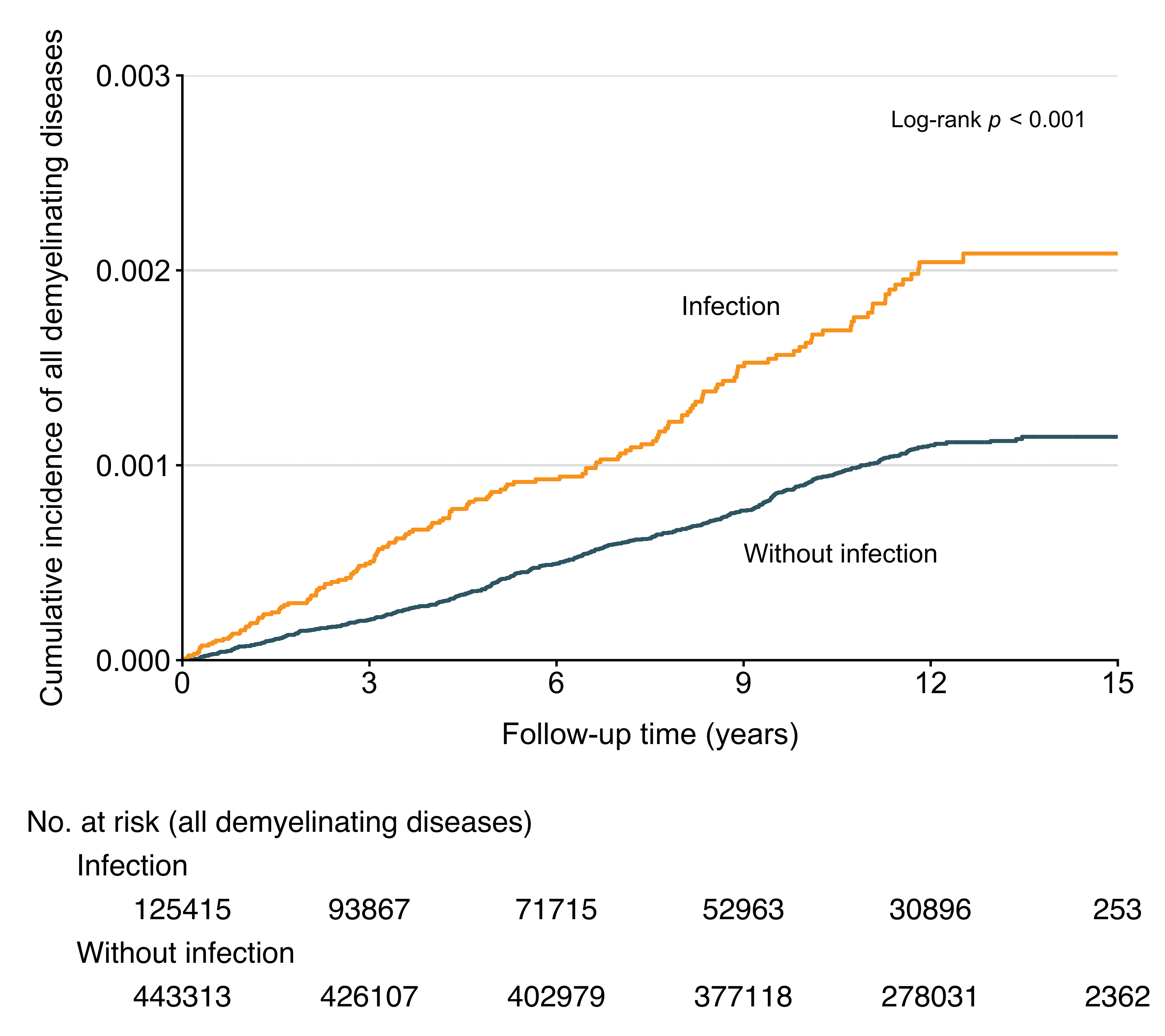

### Figure S2. Cumulative incidence of demyelinating diseases by hospital-treated infectious diseases.

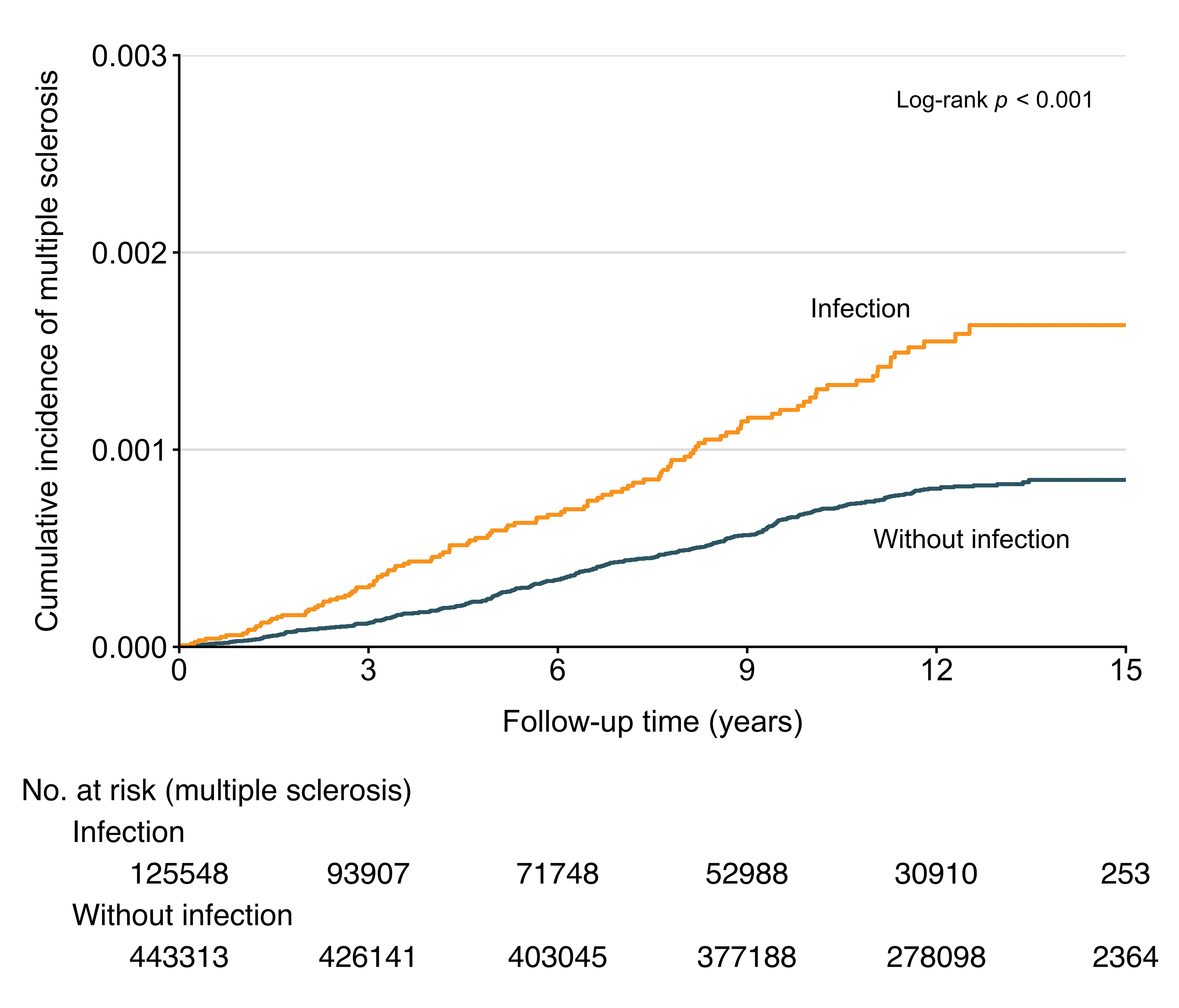

### Figure S3. Cumulative incidence of multiple sclerosis by hospital-treated infectious diseases.

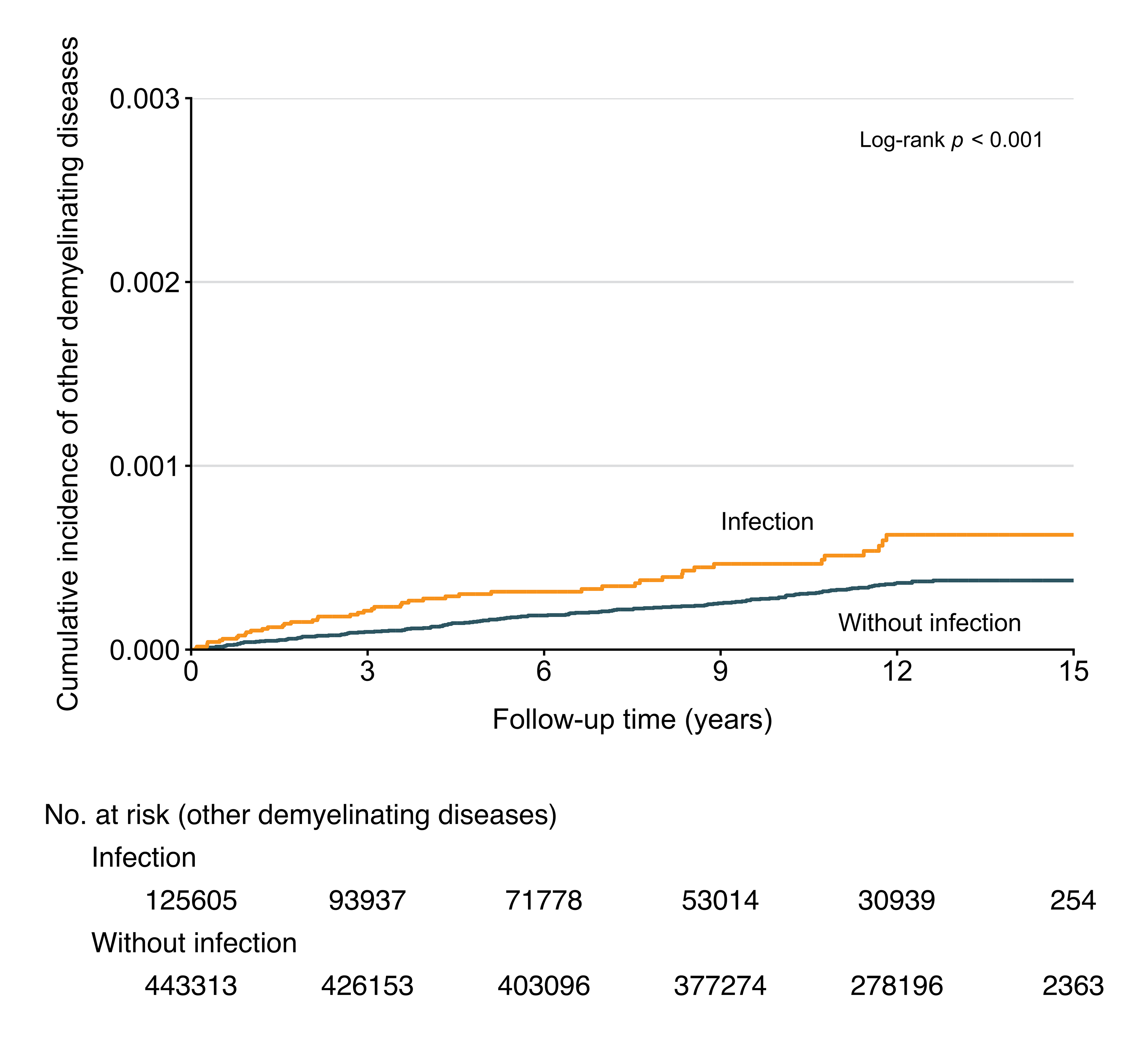

### Figure S4. Cumulative incidence of other demyelinating diseases by hospital-treated infectious diseases.

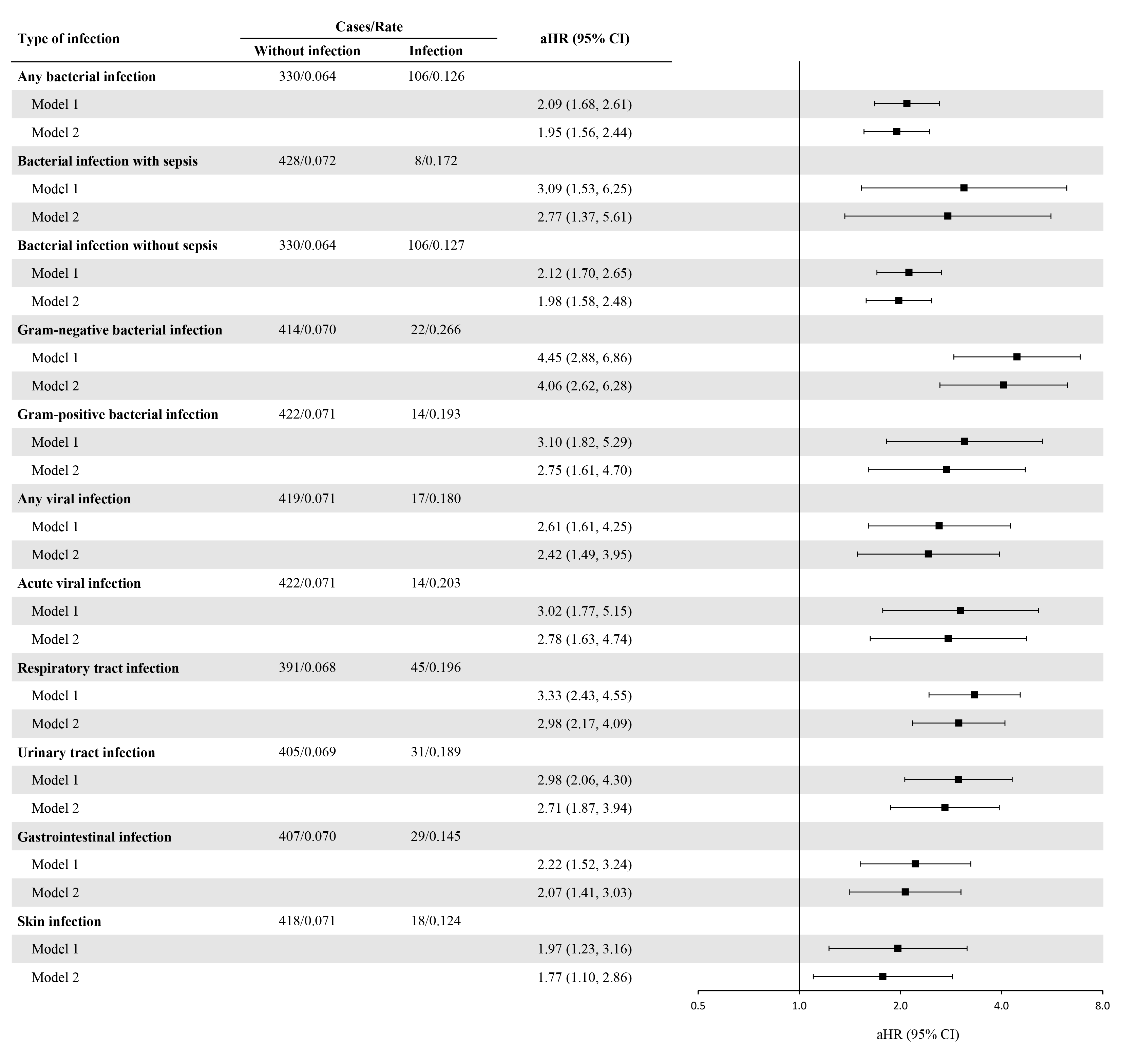

### Figure S5. Risk of multiple sclerosis by type and site of infection.

For each infection type and site, participants without the corresponding infection served as the reference group. Rates are expressed per 1,000 person-years. Model 1 was adjusted for age, sex, education, and Townsend Deprivation Index. Model 2 was additionally adjusted for ethnicity, smoking, alcohol consumption, physical activity, BMI, hypertension, diabetes, cancer, and cardiovascular disease. Abbreviations: aHR, adjusted hazard ratio; BMI, body mass index.

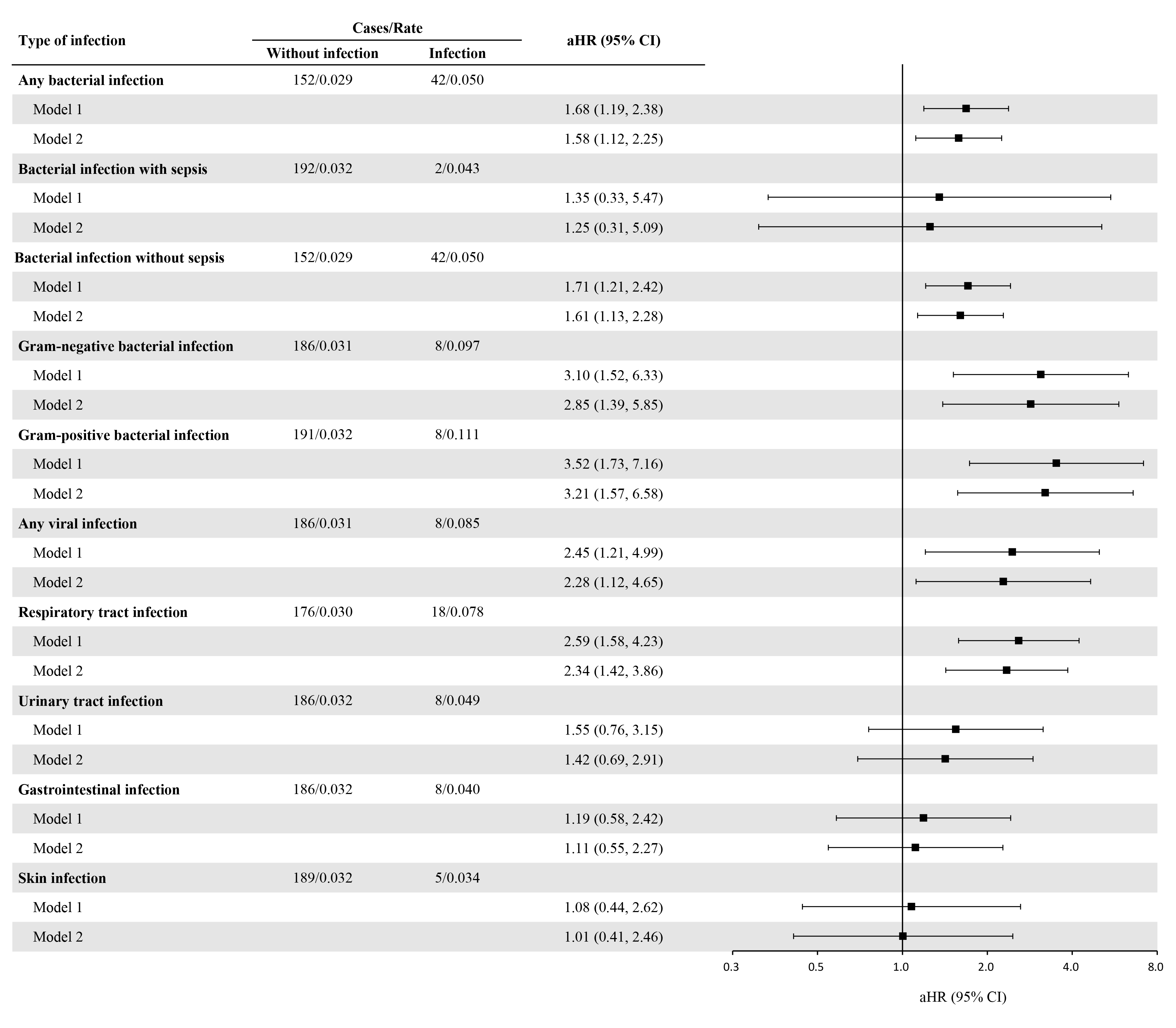

### Figure S6. Risk of other demyelinating diseases by type and site of infection.

For each infection type and site, participants without the corresponding infection served as the reference group. Model 1 was adjusted for age, sex, education, and Townsend Deprivation Index. Model 2 was additionally adjusted for ethnicity, smoking, alcohol consumption, physical activity, BMI, hypertension, diabetes, cancer, and cardiovascular disease. Abbreviations: aHR, adjusted hazard ratio; BMI, body mass index.

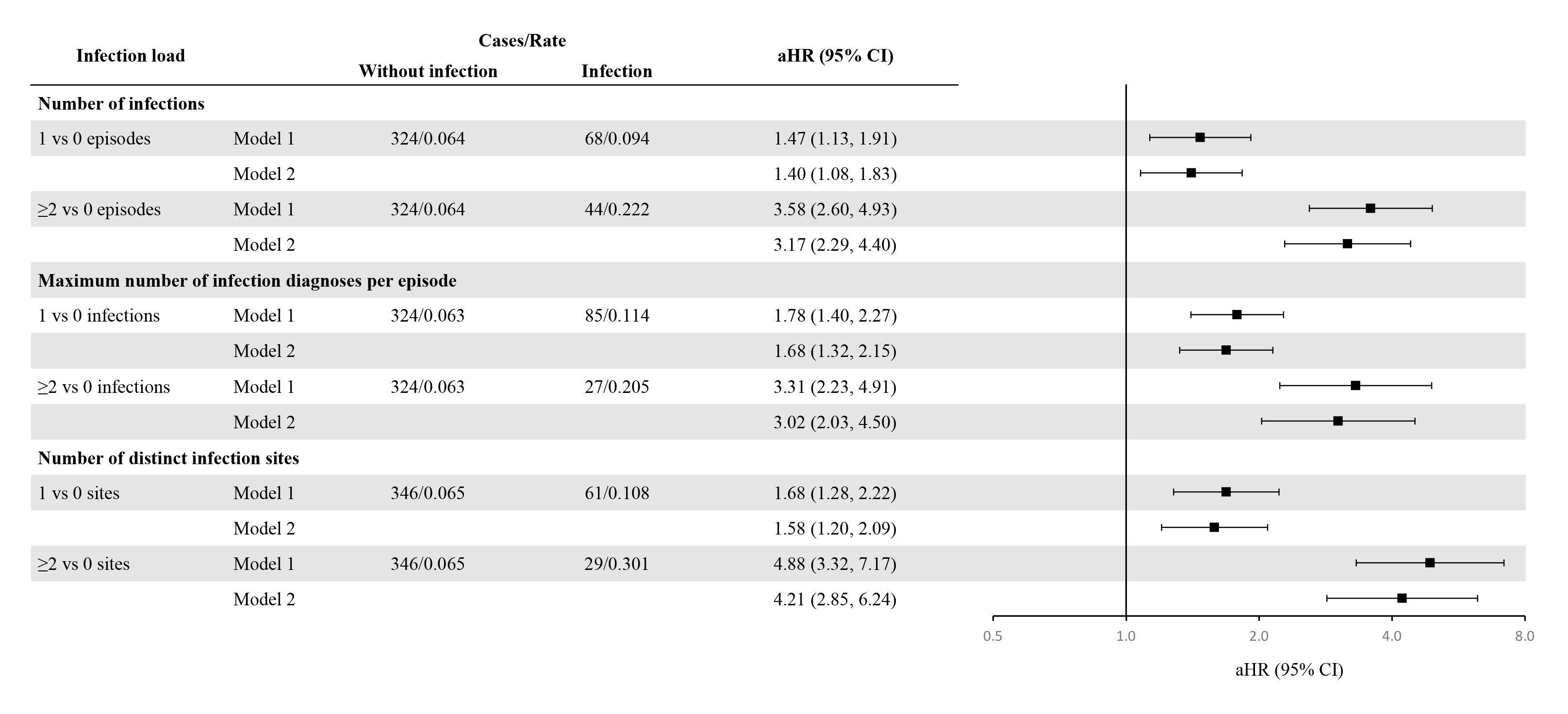

### Figure S7. Infection burden and risk of multiple sclerosis.

aHRs compare participants in each infection burden category with those without infection. Rates are expressed per 1,000 person-years. All *P* for trend < 0.001. Model 1 was adjusted for age, sex, education, and Townsend Deprivation Index. Model 2 was additionally adjusted for ethnicity, smoking, alcohol consumption, physical activity, BMI, hypertension, diabetes, cancer, and cardiovascular disease. Abbreviations: aHR, adjusted hazard ratio; BMI, body mass index.

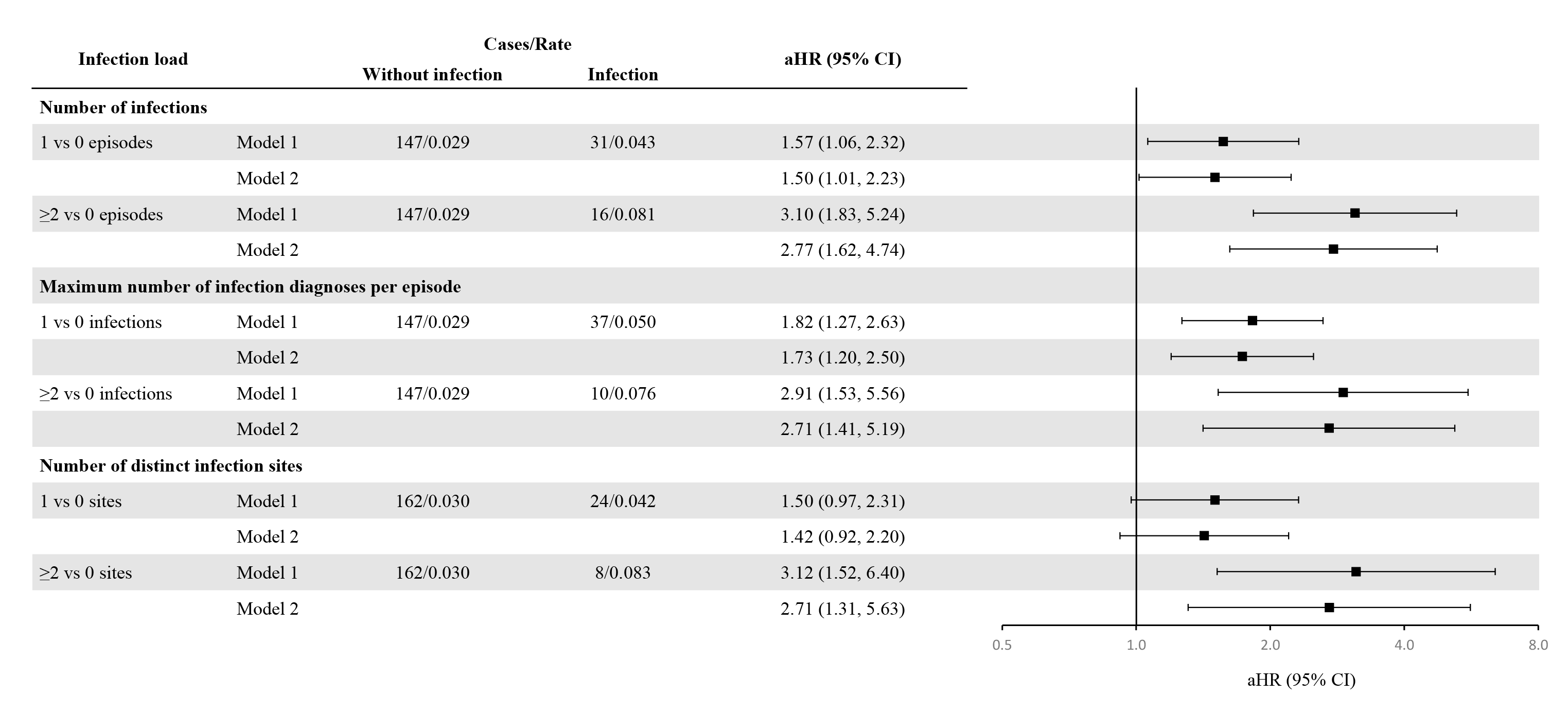

### Figure S8. Infection burden and risk of other demyelinating diseases.

aHRs compare participants in each infection burden category with those without infection. Rates are expressed per 1,000 person-years. *P* for trend for number of infection sites = 0.004, and others < 0.001. Model 1 was adjusted for age, sex, education, and Townsend Deprivation Index. Model 2 was additionally adjusted for ethnicity, smoking, alcohol consumption, physical activity, BMI, hypertension, diabetes, cancer, and cardiovascular disease. Abbreviations: aHR, adjusted hazard ratio; BMI, body mass index.

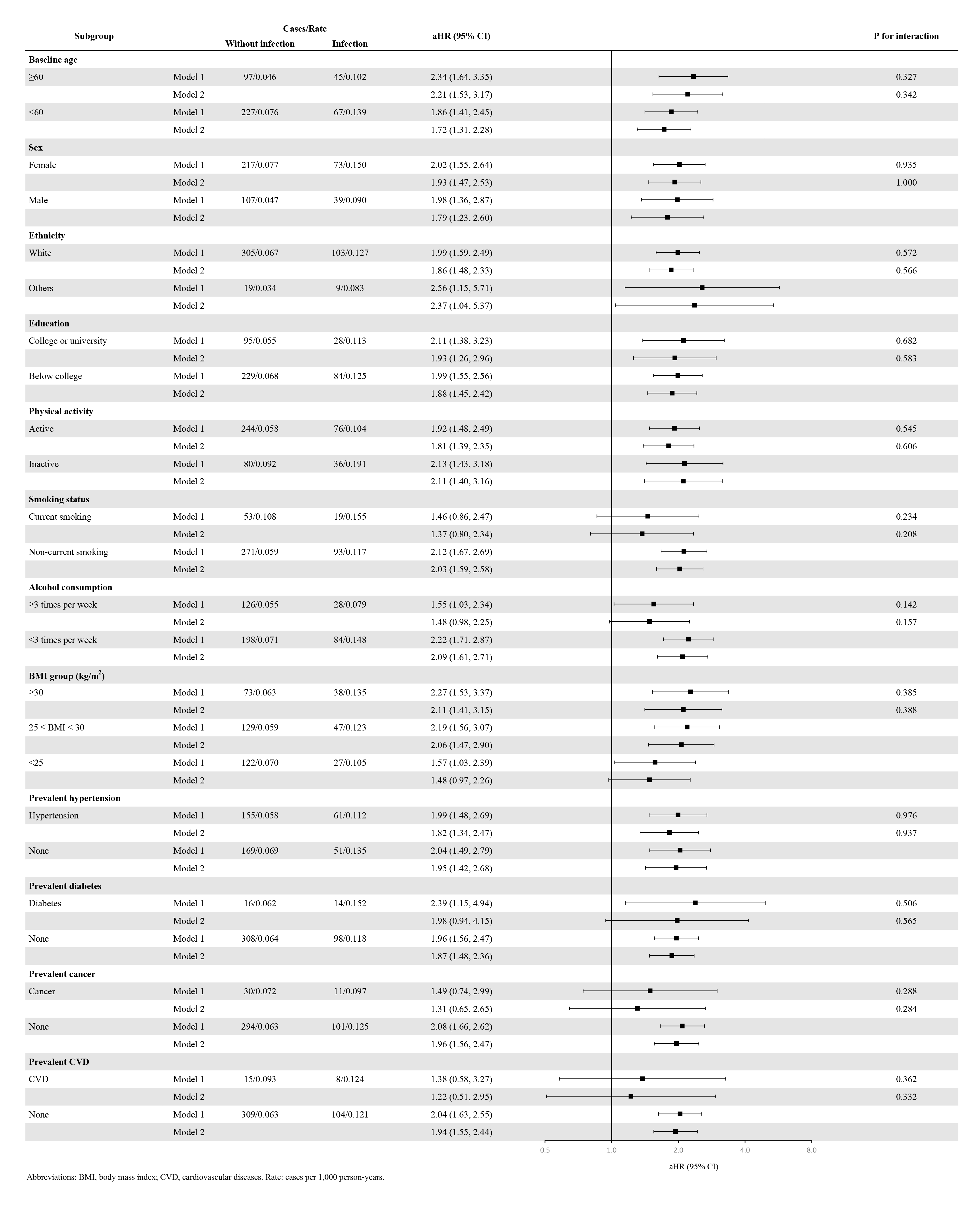

### Figure S9. Associations of hospital-treated infections with the risk of multiple sclerosis by baseline characteristics.

aHRs compare participants with and without hospital-treated infections, with those without infection serving as the reference group. Rates are expressed per 1,000 person-years. P for interaction indicates tests for differences in associations across subgroups. Model 1 was adjusted for age, sex, education, and Townsend Deprivation Index. Model 2 was additionally adjusted for ethnicity, smoking, alcohol consumption, physical activity, BMI, hypertension, diabetes, cancer, and CVD. Abbreviations: aHR, adjusted hazard ratio; BMI, body mass index; CVD, cardiovascular disease.

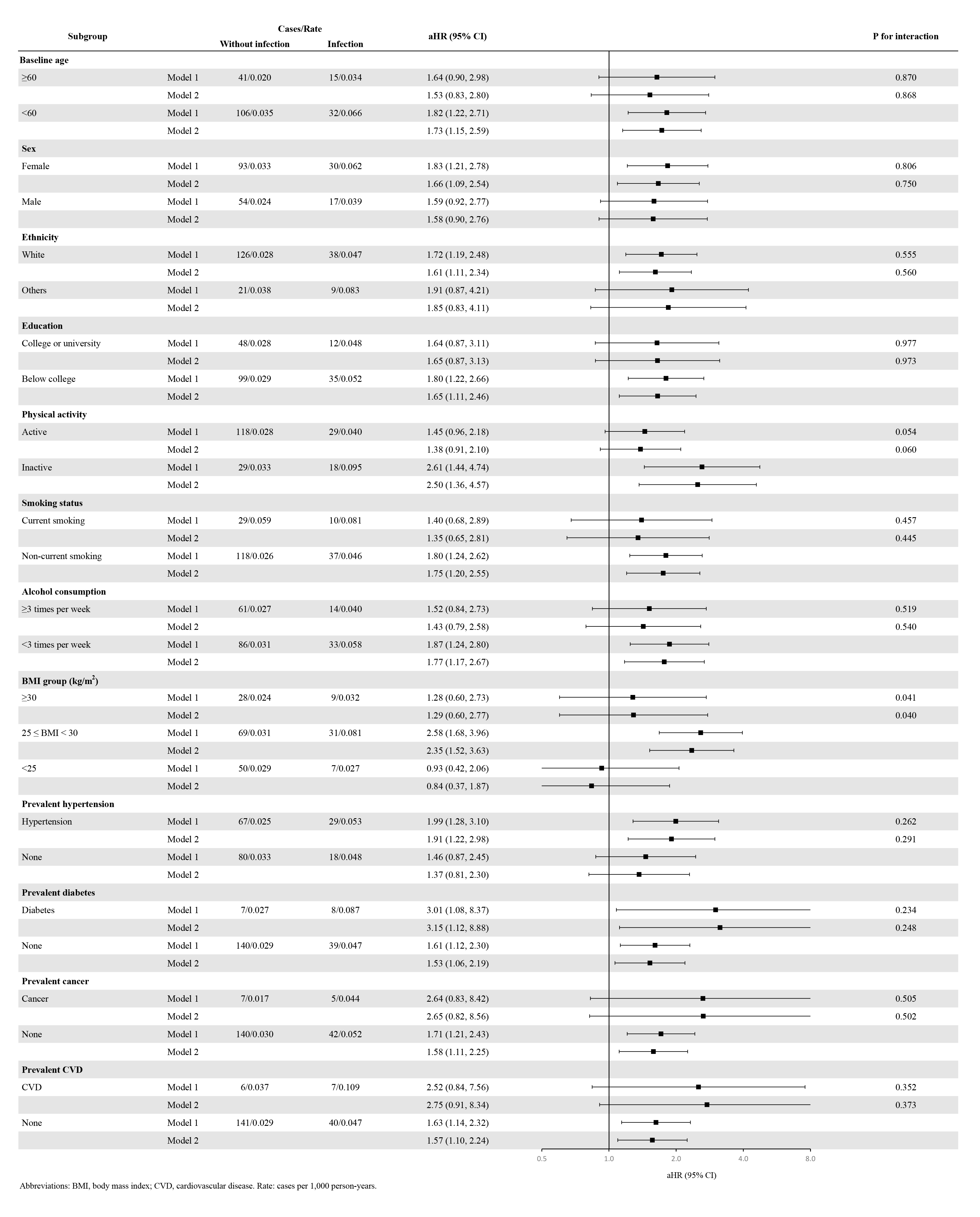

### Figure S10. Associations of hospital-treated infections with the risk of other demyelinating diseases by baseline characteristics.

aHRs compare participants with and without hospital-treated infections, with those without infection serving as the reference group. Rates are expressed per 1,000 person-years. P for interaction indicates tests for differences in associations across subgroups. Model 1 was adjusted for age, sex, education, and Townsend Deprivation Index. Model 2 was additionally adjusted for ethnicity, smoking, alcohol consumption, physical activity, BMI, hypertension, diabetes, cancer, and CVD. Abbreviations: aHR, adjusted hazard ratio; BMI, body mass index; CVD, cardiovascular disease.

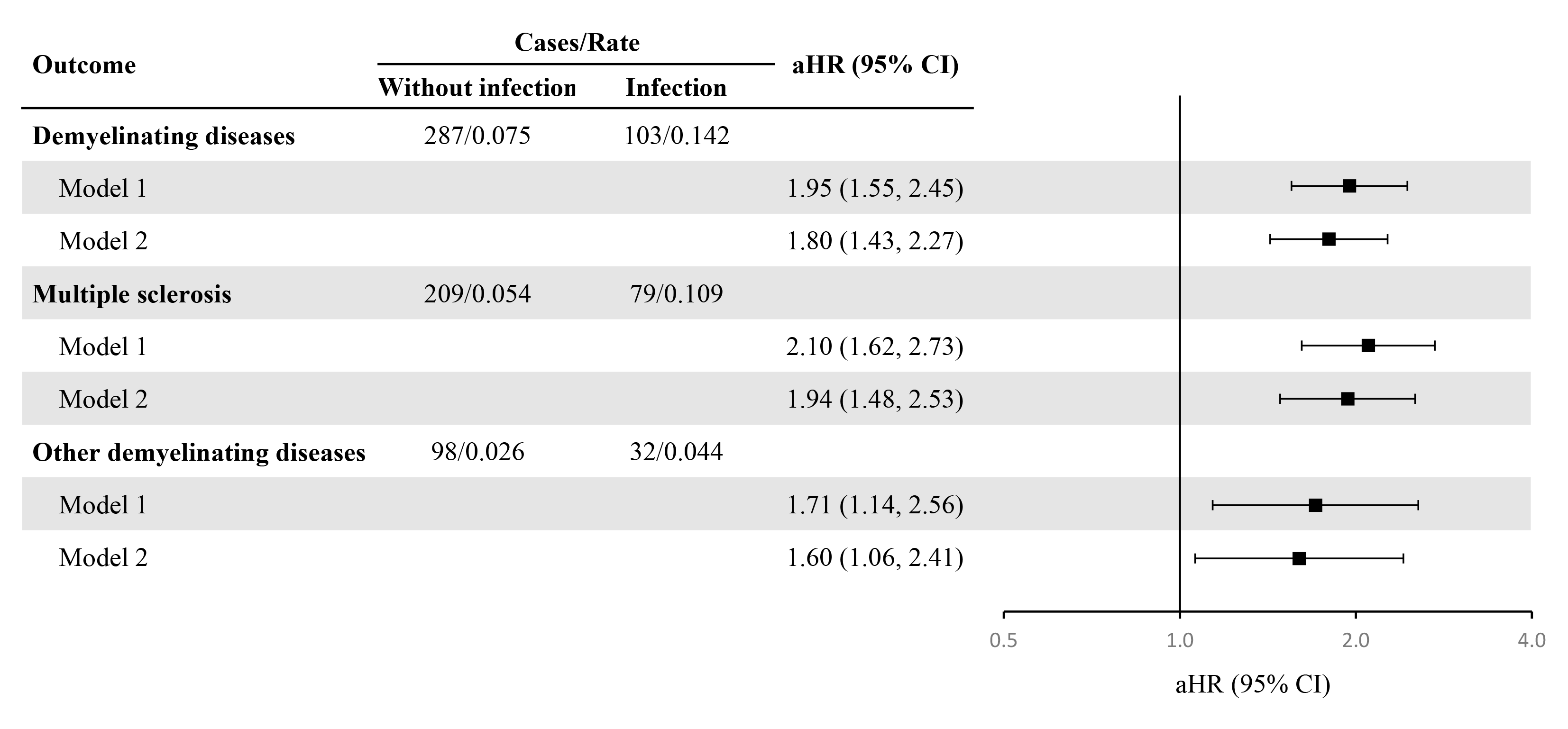

### Figure S11. Associations of hospital-treated infectious diseases with the risk of demyelinating diseases among participants aged ≥50 years at baseline.

Compared with the primary analysis, this restriction excluded 202 cases of demyelinating diseases, 148 cases of MS, and 64 cases of other demyelinating diseases. aHRs compare participants with and without hospital-treated infections, with those without infection serving as the reference group. Rates are expressed per 1,000 person-years. Model 1 was adjusted for age, sex, education, and Townsend Deprivation Index. Model 2 was additionally adjusted for ethnicity, smoking, alcohol consumption, physical activity, BMI, hypertension, diabetes, cancer, and cardiovascular disease. Abbreviations: aHR, adjusted hazard ratio; BMI, body mass index; MS, multiple sclerosis.

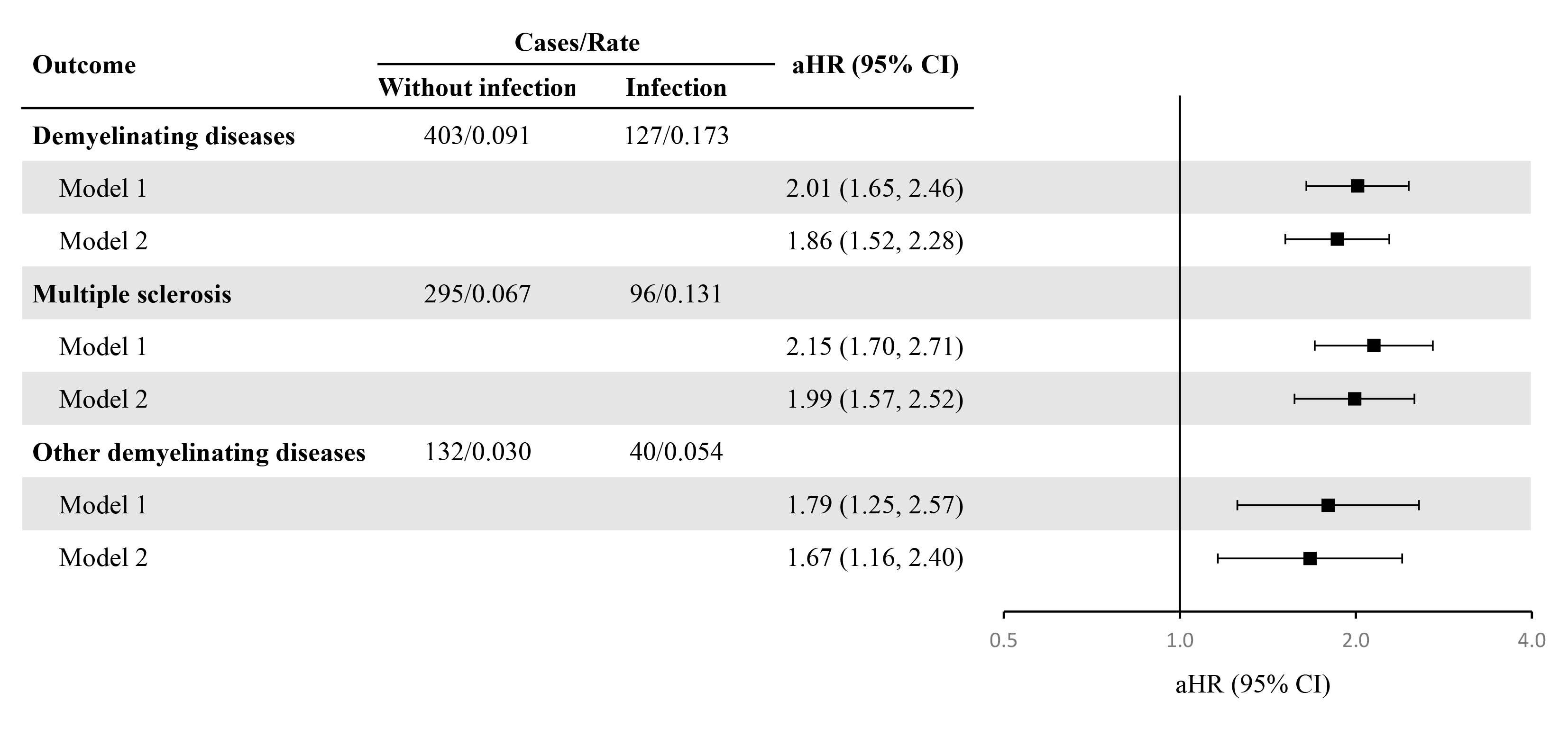

### Figure S12. Associations of hospital-treated infectious diseases with the risk of demyelinating diseases with follow-up censored before the COVID-19 pandemic.

Compared with the primary analysis, censoring follow-up on Dec. 31, 2019 resulted in 62 fewer incident cases of demyelinating diseases, 45 fewer cases of MS, and 22 fewer cases of other demyelinating diseases. aHRs compare participants with and without hospital-treated infections, with those without infection serving as the reference group. Rates are expressed per 1,000 person-years. Model 1 was adjusted for age, sex, education, and Townsend Deprivation Index. Model 2 was additionally adjusted for ethnicity, smoking, alcohol consumption, physical activity, BMI, hypertension, diabetes, cancer, and cardiovascular disease. Abbreviations: aHR, adjusted hazard ratio; BMI, body mass index; MS, multiple sclerosis.

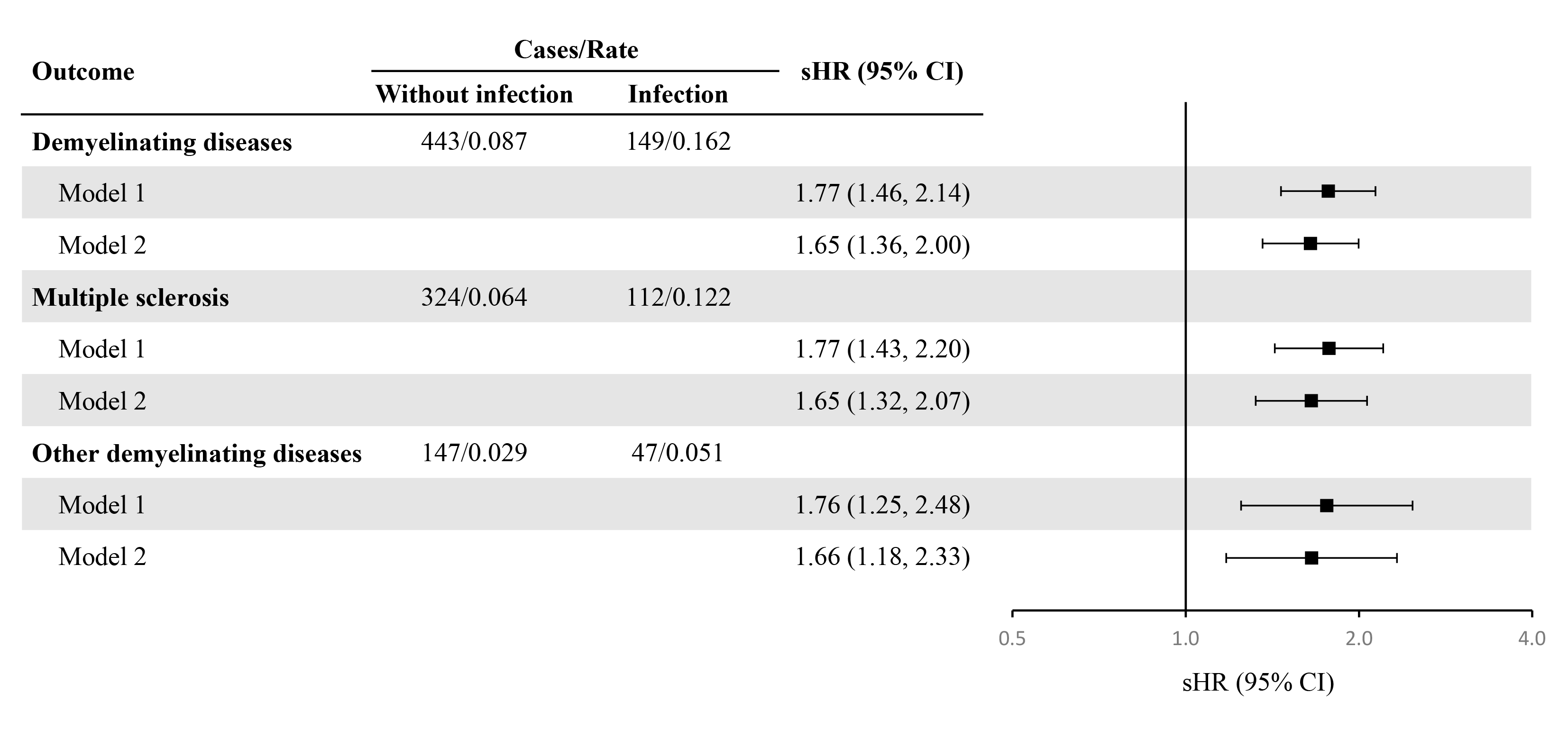

### Figure S13. Associations of hospital-treated infectious diseases with the risk of demyelinating diseases accounting for the competing risk of death.

sHRs compare participants with and without hospital-treated infections, with those without infection serving as the reference group. Model 1 was adjusted for age, sex, education, and Townsend Deprivation Index. Model 2 was additionally adjusted for ethnicity, smoking, alcohol consumption, physical activity, BMI, hypertension, diabetes, cancer, and cardiovascular disease. Abbreviations: BMI, body mass index; sHR, subdistribution hazard ratio.

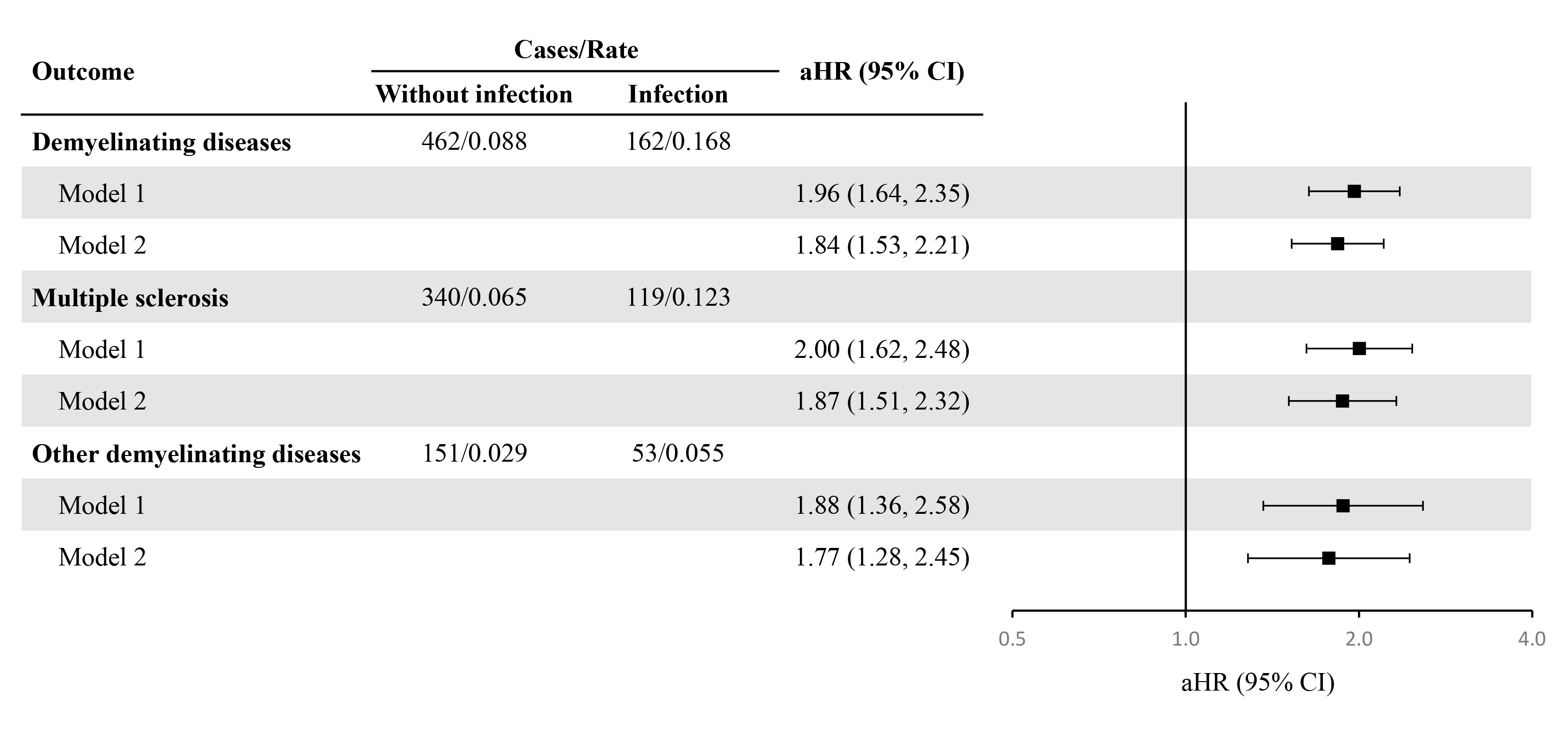

### Figure S14. Associations of hospital-treated infectious diseases with the risk of demyelinating diseases after multiple imputation for missing covariates.

aHRs compare participants with and without hospital-treated infections, with those without infection serving as the reference group. Rates are expressed per 1,000 person-years. Model 1 was adjusted for age, sex, education, and Townsend Deprivation Index. Model 2 was additionally adjusted for ethnicity, smoking, alcohol consumption, physical activity, BMI, hypertension, diabetes, cancer, and cardiovascular disease. Abbreviations: aHR, adjusted hazard ratio; BMI, body mass index.

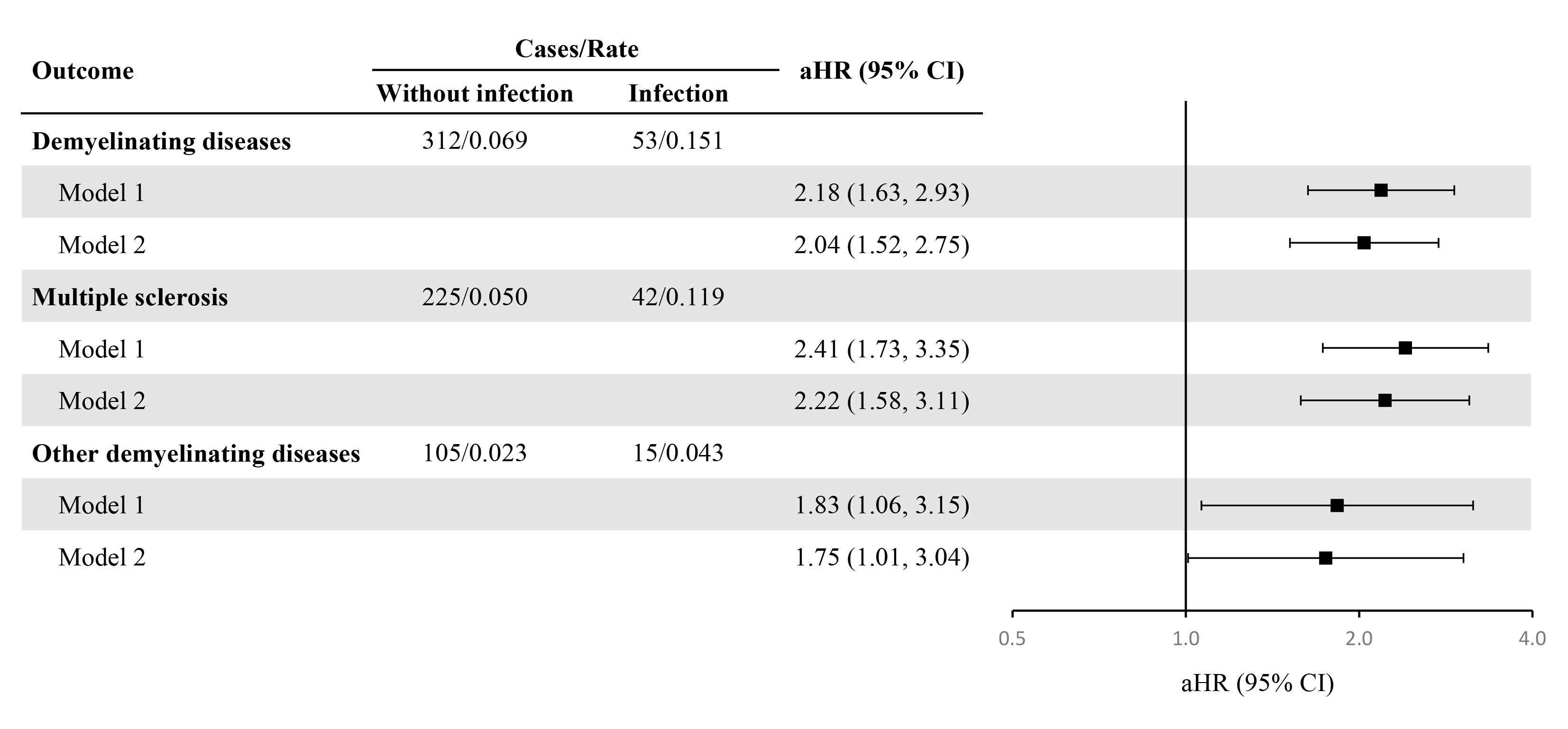

### Figure S15. Associations of hospital-treated infectious diseases with the risk of demyelinating diseases after excluding participants with infections occurring within 3 years before baseline.

Compared with the primary analysis, 227 cases of demyelinating diseases, 169 cases of MS, and 74 cases of other demyelinating diseases were excluded. aHRs compare participants with and without hospital-treated infections, with those without infection serving as the reference group. Rates are expressed per 1,000 person-years. Model 1 was adjusted for age, sex, education, and Townsend Deprivation Index. Model 2 was additionally adjusted for ethnicity, smoking, alcohol consumption, physical activity, BMI, hypertension, diabetes, cancer, and cardiovascular disease. Abbreviations: aHR, adjusted hazard ratio; BMI, body mass index; MS, multiple sclerosis.

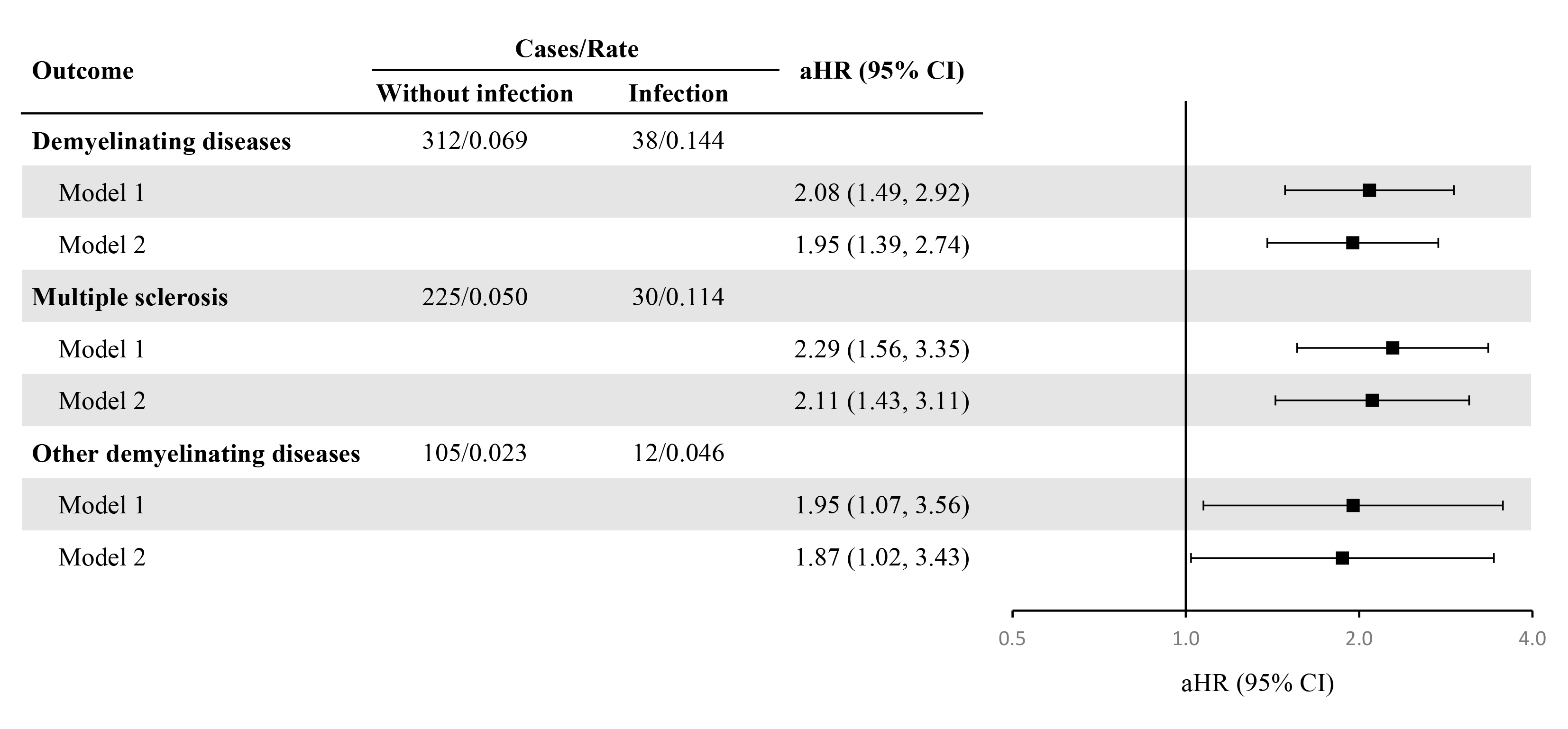

### Figure S16. Associations of hospital-treated infectious diseases with the risk of demyelinating diseases after excluding participants with infections occurring within 5 years before baseline.

Compared with the primary analysis, 242 cases of overall demyelinating diseases, 181 cases of MS, and 77 cases of other demyelinating diseases were excluded. aHRs compare participants with and without hospital-treated infections, with those without infection serving as the reference group. Rates are expressed per 1,000 person-years. Model 1 was adjusted for age, sex, education, and Townsend Deprivation Index. Model 2 was additionally adjusted for ethnicity, smoking, alcohol consumption, physical activity, BMI, hypertension, diabetes, cancer, and cardiovascular disease. Abbreviations: aHR, adjusted hazard ratio; BMI, body mass index; MS, multiple sclerosis.

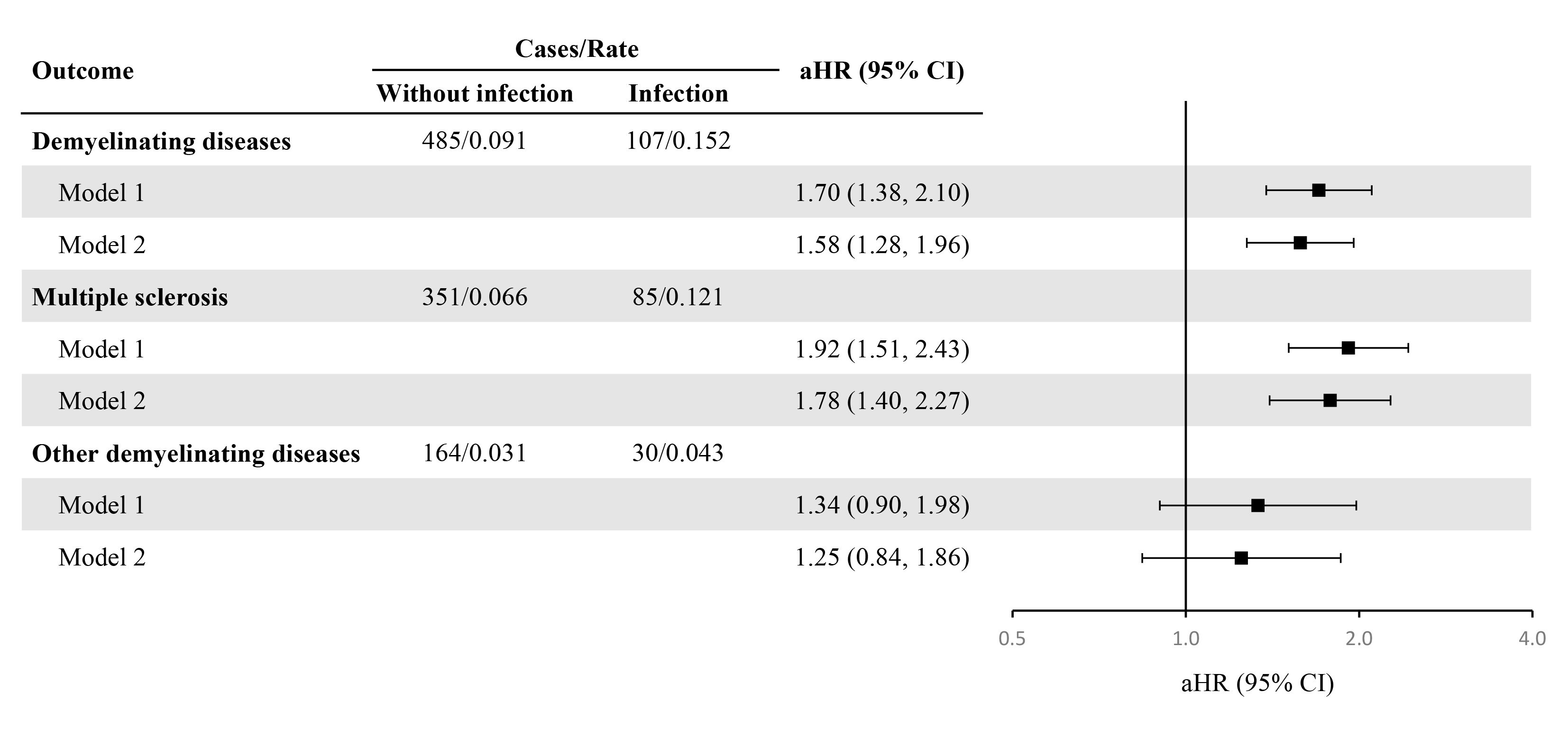

### Figure S17. Associations of hospital-treated infectious diseases with the risk of demyelinating diseases using a 3-year lag period.

42 cases of demyelinating diseases, 27 cases of MS, and 17 cases of other demyelinating diseases were reclassified from the infection group to the without-infection group. aHRs compare participants with and without hospital-treated infections, with those without infection serving as the reference group. Rates are expressed per 1,000 person-years. Model 1 was adjusted for age, sex, education, and Townsend Deprivation Index. Model 2 was additionally adjusted for ethnicity, smoking, alcohol consumption, physical activity, BMI, hypertension, diabetes, cancer, and cardiovascular disease. Abbreviations: aHR, adjusted hazard ratio; BMI, body mass index.

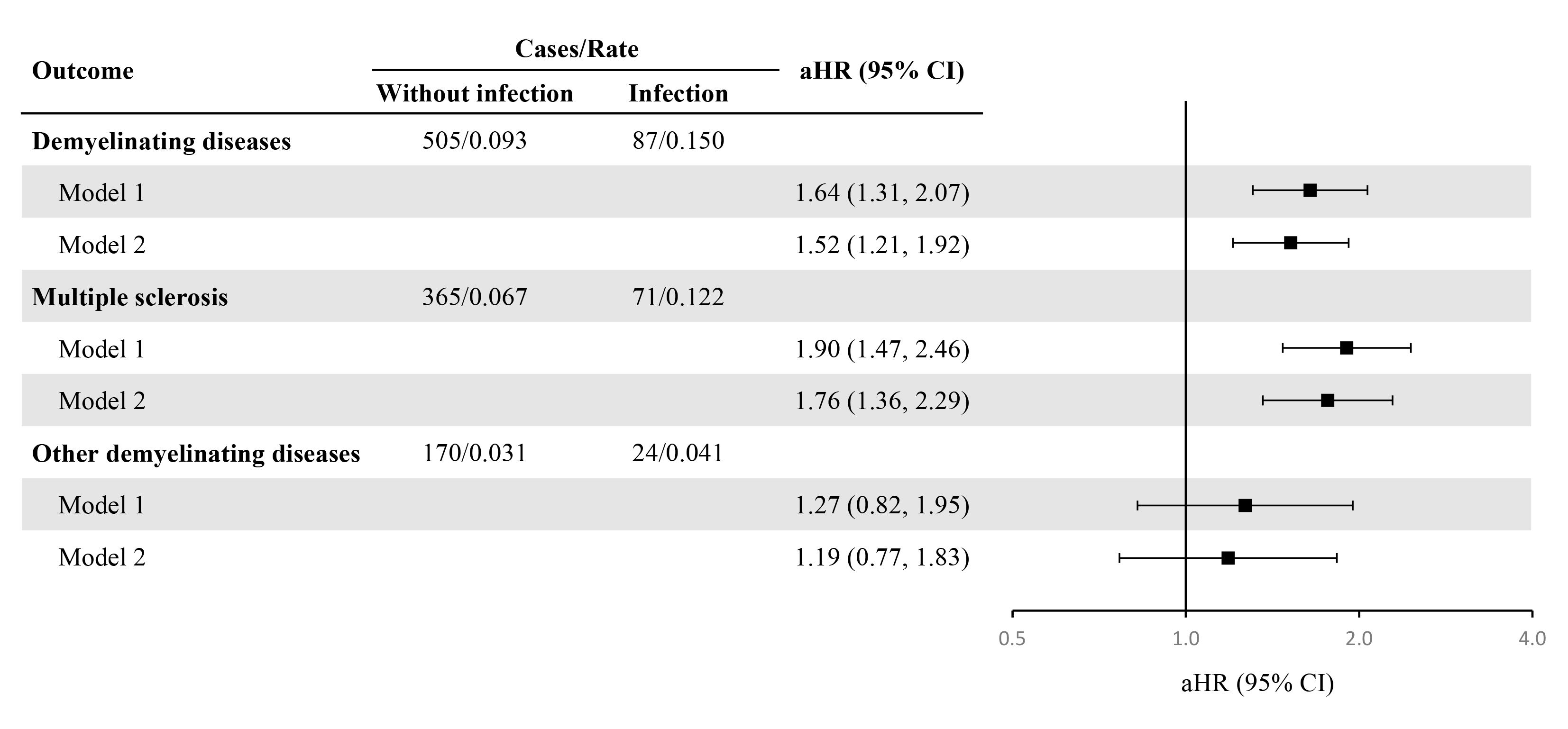

### Figure S18. Associations of hospital-treated infectious diseases with the risk of demyelinating diseases using a 5-year lag period.

**62 cases of demyelinating diseases, 41 cases of MS, and 23 cases of other demyelinating diseases were reclassified from the infection group to the without-infection group.** aHRs compare participants with and without hospital-treated infections, with those without infection serving as the reference group. Rates are expressed per 1,000 person-years. Model 1 was adjusted for age, sex, education, and Townsend Deprivation Index. Model 2 was additionally adjusted for ethnicity, smoking, alcohol consumption, physical activity, BMI, hypertension, diabetes, cancer, and cardiovascular disease. Abbreviations: aHR, adjusted hazard ratio; BMI, body mass index.

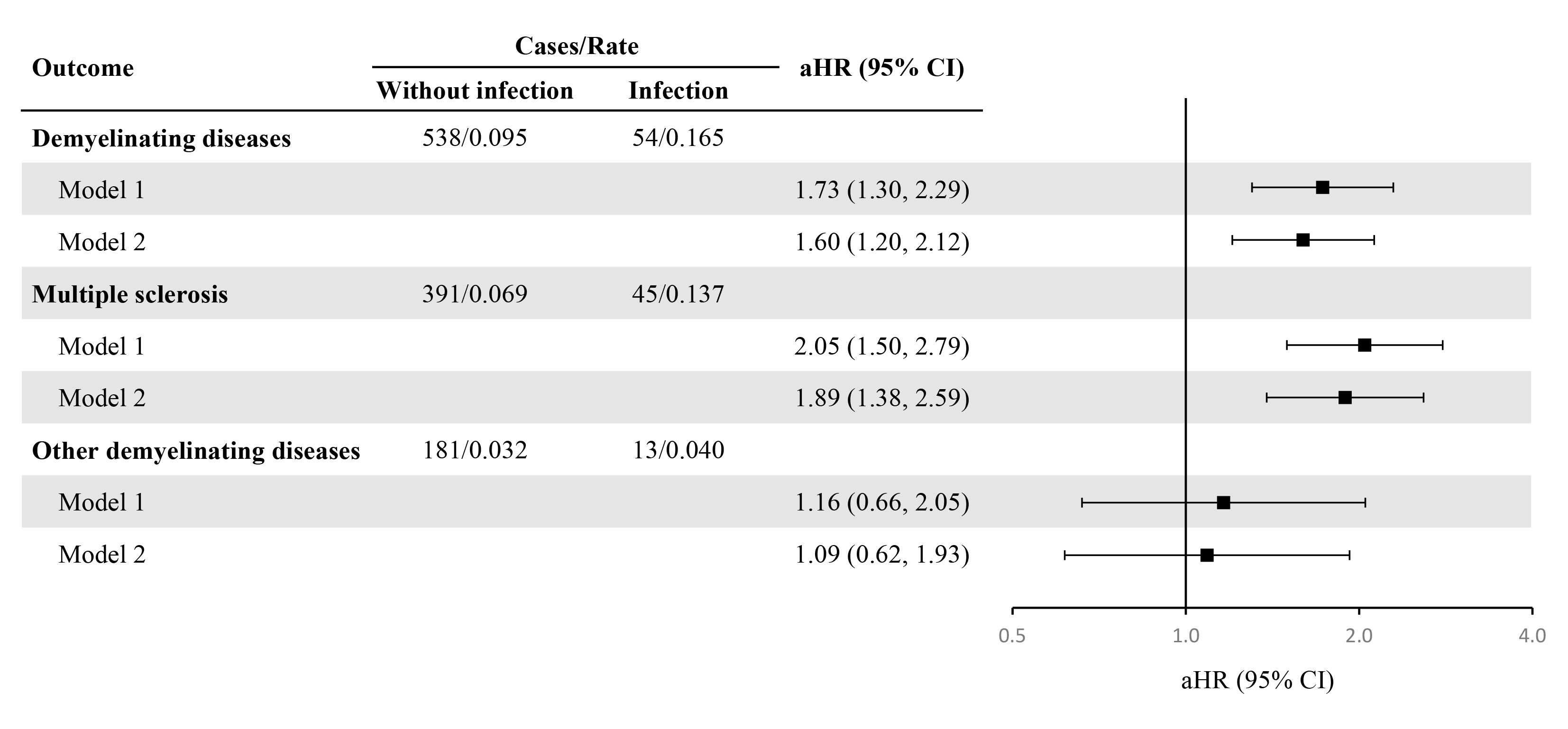

### Figure S19. Associations of hospital-treated infectious diseases with the risk of demyelinating diseases using a 10-year lag period.

95 cases of demyelinating diseases, 67 cases of MS, and 34 cases of other demyelinating diseases were reclassified from the infection group to the without-infection group. aHRs compare participants with and without hospital-treated infections, with those without infection serving as the reference group. Rates are expressed per 1,000 person-years. Model 1 was adjusted for age, sex, education, and Townsend Deprivation Index. Model 2 was additionally adjusted for ethnicity, smoking, alcohol consumption, physical activity, BMI, hypertension, diabetes, cancer, and cardiovascular disease. Abbreviations: aHR, adjusted hazard ratio; BMI, body mass index.
